# Does a prenatal consultation dedicated to fathers’ health widen men’s access to prevention and care? A monocentric interventional research in the Paris metropolitan area

**DOI:** 10.1101/2025.03.13.25323771

**Authors:** Pauline Penot, Gaëlle Jacob, Audrey Guerizec, Valérie-Anne Letembet, Raya Harich, Anne Simon, Miguel de Sousa Mendes, Pierre-Etienne Manuellan, Bruno Renevier, Yazdan Yazdanpanah, Annabel Desgrées du Loû, the Partage Study Group

**Author notes:** Corresponding author: Pauline Penot, 56 bd de la Boissière 93100 Montreuil, France. Membership for the PARTAGE Study Group is provided in the Aknowledgments section.

## Abstract

**Background and objectives:** Prenatal care provides pregnant women with regular opportunities to meet healthcare professionals while not being ill. Nothing similar currently takes place for the second parent. In the PARTAGE project, a prenatal consultation dedicated to future fathers, focused on preserving and improving their own health, has been tested and proven highly acceptable. In this work we analyse the results of this consultation on fathers’ health and to whom it was most useful.

**Design:** PARTAGE was a monocentric interventional study without control arm.

**Setting:** A large public maternity ward in a northeastern suburb of Paris, an area where immigration prevails.

**Participants:** Among 2516 eligible future fathers with effective contact, 1333 were included; fourteen additional sought the consultation directly.

**Intervention:** PARTAGE ran from January 2021 to April 2022 (15 months). A physician or a midwife held a medical interview, extended by a physical examination if symptoms were reported, prescribed immediate biological tests, administered vaccination catch-up when necessary, and offered healthcare and social referral depending on participants’ needs.

**Outcome measures:** In this work we describe the consultation’s effects among participant future fathers: HIV lifetime screening rate, HIV prenatal screening rate and test acceptance among delayed participants; Diphtheria-Tetanus-Pertussis-Poliomyelitis (DTaP-IPV) and Measles-Mumps-Rubella (MMR) vaccination coverage and on-site catch-up rates; diagnosis rate for any medical pathology, including reintegration into care of a lost-to-follow-up pathology; medical and social reference rates.

**Results:** Paternal prenatal consultations resulted in prevention, diagnosis and linkage into care: 37% of participants had never been tested for HIV; 96% had not been tested during or immediately prior to the current pregnancy, of whom 98% accepted HIV screening; 72% were out-of-date for DTaP-IPV or MMR vaccinations, of whom 60% received an on-site catch-up. In addition, 18% had at least one pathology diagnosed or brought back into care; 17% were referred to healthcare professionals and 11% to social workers. Whatever the outcome considered, paternal prenatal consultation benefited socially disadvantaged participants the most: in multivariate analysis, being born in Sub Saharan Africa rather than in France and having no health insurance coverage rather than a complete one were associated with a medical diagnosis (OR 2.67 [1.80-3.98] and OR 2.58 [1.63-4.09], respectively) and with a referral to a healthcare professional (OR 2.67 [1.80-3.98] and OR 3.63 [2.27-5.80], respectively).

**Conclusion:** Paternal prenatal consultations led to the diagnosis of undetected illnesses and to the resumption of care for pathologies that had already been diagnosed but were previously lost to follow-up, especially in the most socially disadvantaged future fathers, who were also enabled to meet social workers when necessary. Paternal prenatal consultations were also broadly useful for catching-up on HIV screening and vaccinations in all expectant fathers. Addressing men’s health during their partner’s pregnancy could help reduce gender and social inequalities in health.

**Clinical Trial number:** NCT05085717; Registry: ClinicalTrials.gov, US National Library of Medicine https://classic.clinicaltrials.gov/ct2/show/NCT05085717. Protocol is available as Supplementary file 1.

**Strengths and limitations of this study:** *Strengths:*

- PARTAGE is, to our knowledge, the first structured health intervention built to address adult men’s health, by using the symbolic event of expecting a child;
- We demonstrated the feasibility and the positive effects of a paternal prenatal consultation;
- In our context, where poverty and immigration prevail, a greater effect was observed among the most disadvantaged men, whose health needs were not met.

*Limitations:*

- In the absence of a control arm, we showed the effects of such consultation, but not a measurable impact.
- Our proactive approach is not directly transferable to routine care. Further research is needed to study how to implement this consultation at a large scale.

## Introduction

### Little is known about future fathers’ health needs, as few health interventions target them

Wherever prenatal care is organized and accessible, women meet healthcare professionals on a regular basis during pregnancy [1]. Prenatal care focuses worldwide on women only. Future fathers are neither patients nor visitors during pregnancy [2]. Consequently, little is known about their health needs, besides the rising description of perinatal depression in men [3] [4]. An Australian “cocooning” program funding Diphtheria-Tetanus-Pertussis-Poliomyelitis (DTaP-IPV) booster for both parents showed better uptake in mothers than in fathers (70% *versus* 53%) [5].

While screening rate for transmissible infections, vaccination, health insurance coverage and HIV, hepatitis B, syphilis or diabetes prevalence are all documented in pregnant women, data on future fathers must be extrapolated from that, scarce, of the general adult male population.

### Gender norms generate health inequalities, and are sustained by healthcare systems

In many contexts, men enjoy more privileges and power than women, but they tend to be in worse health conditions [6]. Men’s poor health reflects several factors, including higher levels of occupational exposure to injury and risk-taking behaviours related to masculinity. Gender norms also shape attitudes to health: men are less likely to visit a doctor when they are ill, or even to report on symptoms [7]. In France, median time from HIV infection to diagnosis was, in 2014, significantly longer in heterosexual men than in heterosexual women, whether among French-born (4.6 *versus* 3 years) or among immigrants (4.5 *versus* 3.1 years) [8]. Given that prenatal HIV screening for women is almost universal in France [9], the absence of a similarly systematic offer targeted at heterosexual men may contribute to this disparity. Health systems contribute to gender inequalities in health, for instance by excluding men from pediatric and prenatal care despite evidence of the benefits of their involvement, perpetuating the stereotype that children’s health is solely the responsibility of women [10].

Social vulnerability combines to gender inequalities in health, and pregnancy is an opportunity to address both.

- *The northeastern suburb of Paris’ specific context*

Seine Saint-Denis (population: 1.7 million inhabitants), located in the northeastern suburbs of Paris, is the department with the highest rates of immigration (31.1%), poverty (30% in 30-50 years adults) and < 1 year mortality in mainland France [11] [12].

Undocumented women are more often covered than undocumented men by the State Medical Assistance (SMA), a specific healthcare benefit that allows eligible illegal immigrants to have free access to most of the French healthcare system [13] [14].

Given that social deprivation worsens men’s health [15], offering men effective access to the healthcare system is necessary, particularly in the most disadvantaged areas.

### A prenatal consultation targeting future fathers

Montreuil city (111 000 inhabitants) hospital provides prenatal care to some 3,500 women every year. A prenatal consultation targeting all future fathers was set up and evaluated in this hospital between 2021 and 2022. We have shown that such consultation was well accepted when it was for the first time organized and actively promoted [16]. In this work, we examine the effects of this consultation on the men who attended it, in terms of prevention (screening and vaccination), diagnosis and linkage with care or social support. We assess the socio-demographic factors associated with each result. We share the experience of a male prenatal consultation and the outcomes of that experience.

## Methods

ANRS PARTAGE (« Prévention, Accès aux droits, Rattrapage vaccinal, Traitement des Affections pendant la Grossesse et pour l’Enfant ») was a monocentric interventional research project which lasted 15 months (January 2021-April 2022) assessing a prenatal consultation offered via the pregnant women to all future fathers in Montreuil hospital, aiming at preserving and improving their own health [16].

### Intervention

Eligible women (18 years of age and older, attending a first prenatal consultation at Montreuil hospital during the study period, reporting a male partner positively involved in the pregnancy and currently living in the Paris metropolitan area) were asked to respond to a face-to-face questionnaire that included their sociodemographic and marital characteristics. Women who consented to having their partner directly offered a consultation provided their partner’s contact information (supplementary file 2). An email was sent to men for whom an email address was provided, with an online appointment platform link, a dedicated phone-line and an email address to schedule a consultation. Fathers who did not reply or who could not be reached by email were called by a research midwife and offered an appointment either during the day or in the evening, on weekdays or Saturday, at the hospital or at the city centre. Participants were asked to bring any vaccination record in their possession.

Consultations, scheduled to last for 30 minutes, were run by a physician or a midwife, who collected consent and the answers to the paternal questionnaire (supplementary file 3), handled clinical examination in case of symptoms and prescribed immediate biological tests (supplementary file 4). Whenever needed, vaccination update was offered: a prescription was issued for pharmacy delivery for fathers with health insurance coverage. The project also had a limited vaccine stock for participants with difficulties to access healthcare. Depending on their choice, men were delivered their test results either face to face, via telephone or via email. The project was backed up by the hospital’s sexual health clinic (CeGIDD), where care is free of charge and where men, depending on their needs, could meet psychologists, physicians, osteopaths and health mediators. An appointment with a social worker was always offered to fathers lacking health insurance coverage, to assist them in acquiring one. Participants could also be referred to any hospital specialist. A local network of partner general practitioners (GPs) was set for participants with none. All participants gave informed consent. Detailed methodology, results on women’s acceptance for participating and men’s uptake of this new offer have been previously described [16].

### Ethics

The French Data Protection Authority (CNIL, registration number 921135) and the French Personal Data Protection Committee (Comité de Protection des Personnes, registration number 21.01.19.44753) gave full ethical approval.

### Statistical analysis

Data on men’s characteristics (country of birth, age, educational level, occupation, employment, previous children, duration and type of union) and on men’s health (previous screenings, HIV and viral hepatitis status, vaccination coverage, medical follow-up) were collected in the paternal questionnaire by the physician or midwife running the consultation (Supplementary file 1).

In addition to the general sociodemographic data, variables exclusively or particularly related to immigrants (participants born abroad with a foreign nationality at birth) were collected: main reason for migration, duration of stay, residence permit, housing conditions, health insurance coverage.

Participants were considered as socially disadvantaged when they had at least one condition from among: lack of stable residence permit, lack of stable housing, lack of complete health insurance coverage, a precarious job or unemployment.

The consultation-related health effects assessed were: medical diagnosis, HIV screening, vaccination update, and health professional and social support linkage. We used the following indicators:

Indicator 1: diagnosis of any medical pathology, or resumption of care of a pathology for which follow-up had been disrupted for at least 2 years;

Indicator 2: proportion of participants accepting HIV testing, in the absence of previous test during the current pregnancy;

Indicator 3: injection of at least one vaccine from the French vaccination schedule;

Indicator 4: referral to at least one healthcare professional;

Indicator 5: referral to a social worker or health mediator.

We studied by univariate analysis the association of indicators 1, 3 and 4 with fathers’ migration status, region of birth, age, level of education, administrative status, duration of residence (for immigrants), employment, health insurance coverage and integration into the healthcare system. All tests were two-sided with p-values < 0.05 defined as statistically significant. Given the limited number of missing data, no imputation was performed. Multivariate models were built to identify sociodemographic factors associated with each indicator after adjustment. The variables included in these models were: region of birth, academic level, age, employment status and health insurance coverage. Duration of stay and administrative status were not included, as they only applied to immigrants. Integration into the healthcare system was not included because of collinearity with health insurance coverage. Migration status was not included for collinearity with region of birth. Backward elimination procedures were used to determine the final models. Analyses were performed specifically on the subpopulation of immigrant men to provide a sensitivity analysis (see supplementary file 7). All analyses were performed in Stata SE 17 (Stata Corporation, College Station, TX, USA).

## Results

### Study population (*Table 1*)

Among 2516 eligible men with effective contact, 1333 were included; fourteen additional future fathers sought the consultation directly (Supplementary file 2). Their median age was 35, interquartile range (IQR) 31-40 years; 40% were expecting their first child; 14% had never been to school, or only to primary school and nearly half had stopped at secondary school level. One-third had precarious working conditions or were unemployed; one-fourth had no GP and had rarely or never been in contact with the French healthcare system. Two-thirds were out of date for Diphtheria-Tetanus-Pertussis-Poliomyelitis (DTaP-IPV) vaccination and 28 % for Measles-Mumps-Rubella (MMR). Lifetime absence of HIV testing was reported by 37% of participants (immigrants: 43%, French born: 29%).

**Table 1:** Characteristics of the study population, Montreuil (France) 2021-2022.

|  | Participants<br>(percentage) | Participants<br>(number) |
| --- | --- | --- |
| **Population size** | 100% | 1347 |
| Of which foreign born | 63% | 842 |
| **Region of birth** |  |  |
| France (mainland and overseas) | 37% | 505 |
| North Africa and Middle East | 18% | 247 |
| Sub-saharan Africa | 29% | 392 |
| Asia | 7% | 96 |
| Rest of Europe, America | 8% | 107 |
| **Age** |  |  |
| < 35 | 49% | 661 |
| 35 and over | 51% | 686 |
| **Education level** |  |  |
| None/Primary or koranic school | 14% | 189 |
| Secondary | 46% | 619 |
| Superior | 40% | 539 |
| **Length of residence in France (for foreign-born)<sup>a</sup>** |  |  |
| >= 7 years | 63% | 526 |
| 3-7 years | 30% | 250 |
| <= 2 years | 8% | 64 |
| **Administrative status** |  |  |
| Legal residency, including French citizenship | 84% | 1129 |
| No residence permit | 13% | 170 |
| < 1 year residence permit | 3% | 48 |
| **Health insurance coverage** |  |  |
| Complete Health Insurance | 74% | 992 |
| Basic Health Insurance | 11% | 155 |
| State Medical Assistance (SMA) | 7% | 91 |
| No | 8% | 109 |
| **Employment status** |  |  |
| Non-precarious employment | 69% | 931 |
| Precarious employment | 18% | 240 |
| Unemployment or training | 13% | 176 |
| **Integration into the healthcare system** |  |  |
| Primary care physician or chronic disease follow-up | 73% | 983 |
| Rarely or never in contact with a physician | 27% | 364 |
| **Diphtheria-Tetanos-Poliomyelitis-Pertussis vaccine coverage<sup>b</sup>** |  |  |
| Up to date | 26% | 357 |
| Unknown vaccination history | 7% | 89 |
| Delayed | 67% | 901 |
| **Measles-Mumps-Rubella vaccine coverage<sup>b</sup>** |  |  |
| 2 doses of vaccine | 40% | 535 |
| No catch-up indication (born <1980 or history of measles) | 10% | 138 |
| Unknown vaccination history | 22% | 296 |
| Non or incompletely vaccinated | 28% | 378 |
| **Lifetime HIV test<sup>c</sup>** |  |  |
| Never | 37% | 505 |
| One or several tests | 62% | 829 |
| Indirectly (blood donation) only | 1% | 12 |

<sup>a</sup> Missing data for 2 participants<sup>b</sup> According to French National recommendations for 2021 https://sante.gouv.fr/archives/archives-presse/archives-communiques-de-presse/article/calendrier-des-vaccinations-2021<sup>c</sup> Missing data for one participant

Among 842 immigrant participants (63%), of which 63% had lived in France for at least 7 years, 47% were born in Sub-saharan Africa, 29% in North Africa and 11% in Asia. One-fifth were illegal; 13% had no health insurance coverage.

Consultation duration often exceeded the scheduled 30 minutes. Physicians progressively substituted midwives in consultations, as participants often complained of symptoms. Whatever the age categories chosen, paternal age was neither associated with a first diagnosis of a medical condition, the resumption of care of a pathology lost to follow-up, with a healthcare referral, nor with on-site vaccination update.

### Pathologies diagnosed or resumption of care

For 240 participants (18%), the consultation led to at least one medical diagnosis or the resumption of care of a chronic condition that had been without follow-up for 2 years or more (for details, see supplementary file 5). Diagnosis and resumption of care were more frequent in immigrant than in French-born participants (Odds Ratio (OR) 2.18 95% [95% Confidence Interval 1.58-3.01]). Socially disadvantaged participants were more likely to be diagnosed or reintegrated into care, as well as participants with no medical follow-up and those who had not attended higher education. Among immigrants, those who had been in France for 3 to 7 years were more likely to receive a new diagnosis or be reintegrated into care (OR 1.70 [1.19-2.42] than those who had arrived more than 7 years before the study. In multivariate analysis, being born in Sub-saharan Africa rather than in France (adjusted OR (aOR) 2.67 [1.80-3.98]), having a lower level of education compared to a university degree (aOR 1.84 [1.30-2.60], secondary school and (aOR 1.81 [1.15-2.85] for none or primary school) and having no health insurance coverage rather than a complete one (aOR 2.58 [1.63-4.09] remained associated with any diagnosis made or resumption of care of any pathology (table 2).

**Table 2:** Univariate and multivariate logistic regression analysis of factors associated with any diagnosis or pathology brought back into care during paternal prenatal consultation, Montreuil, 2021-2022.

|  | Participants<br>(n/N) | Percentage of<br>participants | Univariate analysis<br>OR [IC 95] N = 1347 | Final multivariate model<sup>a</sup><br>aOR [IC 95] N=1347 |
| --- | --- | --- | --- | --- |
| Participants with any diagnosis or pathology brought back into care | 240/1347 | 18 % |  |  |
| **Migration status** |  |  |  |  |
| French-born | 57/505 | 11 % | 1.00 |  |
| Immigrant | 183/842 | 22 % | **2.18 [1.58-3.01]** |  |
| **Region of birth** |  |  |  |  |
| France | 57/505 | 11 % | 1.00 | 1.00 |
| North Africa-Middle East | 28/247 | 11 % | 1.00 [0.62-1.62] | 0.83 [0.50-1.36] |
| Sub-saharan Africa | 124/392 | 32 % | **3.64 [2.57-5.15]** | **2.67 [1.80-3.98]** |
| Asia | 16/96 | 17% | 1.57 [0.86-2.87] | 1.34 [0.73-2.49] |
| Rest of Europe-America | 15/107 | 14 % | 1.28 [0.69-2.36] | 0.89 [0.47-1.69] |
| **School level** |  |  |  |  |
| University | 61/539 | 11 % | 1.00 | 1.00 |
| Secondary school | 123/619 | 20 % | **1.94 [1.39-2.71]** | **1.84 [1.30-2.60]** |
| None, primary or koranic school | 56/189 | 30 % | **3.22 [2.13-4.85]** | **1.81 [1.15-2.85]** |
| **Age** |  |  |  |  |
| < 35 | 114/661 | 17 % | 1.00 |  |
| 35 and over | 126/686 | 18 % | 1.08 [0.82-1.43] |  |
| **Administrative status** |  |  |  |  |
| Regular situation | 164/1129 | 15 % | 1.00 |  |
| No or < 1 year residence permit | 76/218 | 35 % | **3.15 [2.28-4.35]** |  |
| **Length of stay (immigrants)** |  |  |  |  |
| 7 years and over | 98/526 | 18 % | 1.00 |  |
| 3-7 years | 70/250 | 28 % | **1.70 [1.19-2.42]** |  |
| <= 2 years | 15/64 | 23 % | 1.34 [0.72-2.48] |  |
| **Employment status** |  |  |  |  |
| Non-precarious employment | 138/931 | 15 % | 1.00 |  |
| Precarious employment | 62/240 | 26 % | **2.00 [1.42-2.81]** |  |
| Unemployment or training | 40/176 | 23 % | **1.69 [1.14-2.51]** |  |
| **Health insurance coverage** |  |  |  |  |
| Complete Health Insurance | 148/992 | 15 % | 1.00 | 1.00 |
| Basic Health Insurance | 25/155 | 16 % | 1.10 [0.70-1.74] | 0.91 [0.56-1.47] |
| State Medical Assistance (SMA) | 22/91 | 24 % | **1.82 [1.10-3.03]** | 1.07 [0.62-1.87] |
| No | 45/109 | 41 % | **4.01 [2.64-6.10]** | **2.58 [1.63-4.09]** |
| **Integration into the healthcare system** |  |  |  |  |
| Primary care physician or chronic disease follow-up | 147/983 | 15 % | 1.00 |  |
| Rarely or never in contact with a physician | 93/364 | 26 % | **1.95 [1.45-2.61]** |  |
| * Variables included in the complete multivariate model were: region of birth, school level, age, employment status, health insurance coverage. Length of stay and administrative status were excluded, as they only applied to immigrants. Integration into the healthcare system was excluded because of collinearity with health insurance coverage. Migration status was excluded for collinearity with region of birth. |  |  |  |  |

### HIV screening acceptance rate

Among 1297 participants (96%) not tested during the current pregnancy or as part of a medically assisted procreation, 1276 (98%) had an HIV test following consultation, without HIV incident discovery. The refusal rate was 1.1% (see Supplementary file 5 for details).

### Referral to healthcare professionals

For 225 participants (17%), consultation resulted in a referral to at least one healthcare professional (for details, see appendix 3). Immigrant participants were more likely than their French-born counterparts to be referred to healthcare professionals (OR 1.86 [1.35-2.56]). Paternal age was not associated with referral. As for diagnosis, socially disadvantaged participants and those who had not attended higher education were more likely to be referred to at least one healthcare professional. In multivariate analysis, being born in Sub-saharan Africa rather than in France (aOR 1.89 [1.26-2.84]), having a secondary level of education rather than a university degree (aOR 1.74 [1.22-2.47]) and having no health insurance coverage rather than a complete one (aOR 3.63 [2.27-5.80] remained associated with referral to one or several healthcare professionals (table 3).

**Table 3:** Univariate and multivariate logistic regression analysis of factors associated with any healthcare professional referral during paternal prenatal consultation, Montreuil, 2021-2022.

|  | Participants<br>(n/N) | Percentage of<br>participants | Univariate analysis<br>OR [IC 95] N = 1347 | Final multivariate model<sup>a</sup><br>aOR [IC 95] N=1347 |
| --- | --- | --- | --- | --- |
| Participants referred to one or several<br>healthcare professionals | 225/1347 | 17 % |  |  |
| **Migration status** |  |  |  |  |
| French-born | 59/505 | 12 % | 1.00 |  |
| Immigrant | 166/842 | 20 % | **1.86 [1.35-2.56]** |  |
| **Region of birth** |  |  |  |  |
| France | 59/505 | 12 % | 1.00 | 1.00 |
| North Africa-Middle East | 25/247 | 10 % | 0.85 [0.52-1.40] | 0.62 [0.37-1.05] |
| Sub-saharan Africa | 108/392 | 28 % | **2.88 [2.02-4.08]** | **1.89 [1.26-2.84]** |
| Asia | 18/96 | 19 % | 1.74 [0.98-3.12] | 1.45 [0.80-2.63] |
| Rest of Europe-America | 15/107 | 14 % | 1.23 [0.67-2.27] | 0.76 [0.40-1.46] |
| **School level** |  |  |  |  |
| University | 59/539 | 11 % | 1.00 | 1.00 |
| Secondary school | 118/619 | 19 % | **1.92 [1.37-2.68]** | **1.74 [1.22-2.47]** |
| None, primary or koranic school | 48/189 | 25 % | **2.77 [1.81-4.23]** | 1.50 [0.93-2.41] |
| **Age** |  |  |  |  |
| < 35 | 112/661 | 17 % | 1.00 |  |
| 35 and over | 113/686 | 16 % | 0.97 [0.73-1.29] |  |
| **Administrative status** |  |  |  |  |
| Regular situation | 153/1129 | 14 % | 1.00 |  |
| No or < 1 year residence permit | 72/218 | 33 % | **3.15 [2.26-4.37]** |  |
| **Length of stay (immigrants)** |  |  |  |  |
| 7 years and over | 84/526 | 16 % | 1.00 |  |
| 3-7 years | 65/250 | 26 % | **1.85 [1.28-2.67]** |  |
| <= 2 years | 17/64 | 27 % | **1.90 [1.04-3.47]** |  |
| **Employment status** |  |  |  |  |
| Non-precarious employment | 127/931 | 14 % | 1.00 |  |
| Precarious employment | 60/240 | 25 % | **2.11 [1.49-2.98]** |  |
| Unemployment or training | 38/176 | 22 % | **1.74 [1.16-2.61]** |  |
| **Health insurance coverage** |  |  |  |  |
| Complete Health Insurance | 129/992 | 13 % | 1.00 | 1.00 |
| Basic Health Insurance | 28/155 | 18 % | 1.47 [0.94-2.31] | 1.30 [0.82-2.07] |
| State Medical Assistance (SMA) | 23/91 | 25 % | **2.26 [1.36-3.76]** | 1.68 [0.96-2.92] |
| No | 45/109 | 41 % | **4.70 [3.08-7.19]** | **3.63 [2.27-5.80]** |
| **Integration into the healthcare system** |  |  |  |  |
| Primary care physician or chronic disease<br>follow-up | 105/983 | 11 % | 1.00 |  |
| Rarely or never in contact with a physician | 120/364 | 33 % | **4.11 [3.05-5.53]** |  |
| <sup>a</sup> Variables included in the complete multivariate model were: region of birth, school level, age, employment status, health insurance coverage. Length of stay and administrative status were excluded, as they only applied to immigrants. Integration into the healthcare system was excluded because of collinearity with health insurance coverage. Migration status was excluded for collinearity with region of birth. |  |  |  |  |

### Vaccination updates

Among all participants, 591 (44%) received on-site vaccination updates. Immigrant participants were more likely to receive on-site vaccination updates than their French-born counterparts (OR 2.39 [1.90-3.02]). Socially disadvantaged participants were more likely to receive on-site vaccination updates, as well as participants with no medical follow-up and those who had not attended higher education. Among immigrants, those living in France for 3 to 7 years were more likely to receive a vaccination update, compared to those living in France for at least 7 years. The same trend emerges for new immigrants but the statistical power is insufficient for the OR to be significant.

In multivariate analysis, being born in Sub-saharan Africa, Asia or North Africa-Middle East rather than in France (aOR 1.99 [1.46-2.72]), 1.83 [1.16-2.88], 1.47 [1.06-2.03], respectively), having primary schooling level or less rather than a university degree (aOR 1.75 [1.20-2.56]) and having either no health insurance coverage (aOR 3.16 [1.98-5.04], SMA (aOR 1.68 [1.05-2.67]) or only basic health insurance (aOR 1.69 [1.19-2.41]) rather than a complete standard health insurance coverage, remained associated with on-site vaccine update (table 4).

**Table 4:** Univariate and multivariate regression analysis of factors associated with vaccine update during paternal prenatal consultation, Montreuil, 2021-2022.

|  | Participants<br>(n/N) | Percentage of<br>participants | Univariate analysis<br>OR [IC 95] N = 1347 | Final multivariate model<sup>a</sup><br>aOR [IC 95] N=1347 |
| --- | --- | --- | --- | --- |
| Participants with one or several vaccine updates | 591/1347 | 44 % |  |  |
| **Migration status** |  |  |  |  |
| French-born | 156/505 | 31 % | 1.00 |  |
| Immigrant | 435/842 | 52 % | **2.39 [1.90-3.02]** |  |
| **Region of birth** |  |  |  |  |
| France | 156/505 | 31 % | 1.00 | 1.00 |
| North Africa-Middle East | 111/247 | 45 % | **1.83 [1.33-2.50]** | **1.47 [1.06-2.03]** |
| Sub-saharan Africa | 226/392 | 58 % | **3.05 [2.31-4.01]** | **1.99 [1.46-2.72]** |
| Asia | 48/96 | 50 % | **2.24 [1.44-3.48]** | **1.83 [1.16-2.88]** |
| Rest of Europe-America | 50/107 | 47 % | **1.96 [1.28-3.00]** | 1.41 [0.90-2.20] |
| **School level** |  |  |  |  |
| University | 197/539 | 37 % | 1.00 | 1.00 |
| Secondary school | 275/619 | 44 % | **1.39 [1.10-1.76]** | 1.19 [0.93-1.53] |
| None, primary or koranic school | 119/189 | 63 % | **2.95 [2.09-4.16]** | **1.75 [1.20-2.56]** |
| **Age** |  |  |  |  |
| < 35 | 273/661 | 41 % | 1.00 |  |
| 35 and over | 318/686 | 46 % | 1.23 [1.00-1.52] |  |
| **Administrative status** |  |  |  |  |
| Regular situation | 444/1129 | 39 % | 1.00 |  |
| No or < 1 year residence permit | 147/218 | 67 % | **3.19 [2.35-4.34]** |  |
| **Length of stay (immigrants)** |  |  |  |  |
| 7 years and over | 252/526 | 48 % | 1.00 |  |
| 3-7 years | 145/250 | 58 % | **1.50 [1.11-2.03]** |  |
| <= 2 years | 37/64 | 58 % | 1.49 [0.88-2.52] |  |
| **Employment status** |  |  |  |  |
| Non-precarious employment | 368/931 | 40 % | 1.00 |  |
| Precarious employment | 136/240 | 57 % | **2.00 [1.50-2.67]** |  |
| Unemployment or training | 87/176 | 49 % | **1.49 [1.08-2.07]** |  |
| **Health insurance coverage** |  |  |  |  |
| Complete Health Insurance | 372/992 | 38 % | 1.00 | 1.00 |
| Basic Health Insurance | 84/155 | 54 % | **1.97 [1.40-2.77]** | **1.69 [1.19-2.41]** |
| State Medical Assistance (SMA) | 55/91 | 60 % | **2.55 [1.64-3.95]** | **1.68 [1.05-2.67]** |
| No | 80/109 | 73 % | **4.60 [2.95-7.17]** | **3.16 [1.98-5.04]** |
| **Integration into the healthcare system** |  |  |  |  |
| Primary care physician or chronic disease follow-up | 393/983 | 40 % | 1.00 |  |
| Rarely or never in contact with a physician | 198/364 | 54 % | **1.79 [1.40-2.28]** |  |

<sup>a</sup> Variables included in the complete multivariate model were: region of birth, school level, age, employment status, health insurance coverage. Length of stay and administrative status were excluded, as they only applied to immigrants. Integration into the healthcare system was excluded because of collinearity with health insurance coverage. Migration status was excluded for collinearity with region of birth.

In addition, 145 participants (11%), most of them being immigrants (142/842, 17% *versus* French-born participants 3/505, 0.6%) were referred to a social worker or a health mediator (see Supplementary file 6).

## Discussion

In this study, paternal prenatal consultation led to the re-engagement of a significant number of future fathers with HIV screening and vaccination, both of which were overdue. Consultation led, for 18% of participants, to a medical diagnosis or to resumption of care for a pathology for which follow-up had been discontinued (for a list of most common diagnosed pathologies, see appendix 2); 17% were referred to one or several healthcare professionals and 11% to a social worker.

37% of all participants had never been tested for HIV, as no systematic offer targets heterosexual men, despite demonstration that maternal seroconversion during pregnancy is responsible for at least 34% of HIV-infected births in the United Kingdom [17]. Pregnancy and childbirth is a good opportunity for heterosexual men to bridge the gender gap in HIV testing. Nonetheless, no new HIV infections were discovered during the study. Besides the low prevalence of undiagnosed HIV in France, one explanation could be that HIV-positive women largely refused (12/14, data not shown) their partner to be contacted, precisely for fear of disclosure of their own HIV infection following a possible HIV diagnosis of their partners. A paternal prenatal consultation offered earlier in pregnancy and starting from the first pregnancy would bring us closer to the World Health Organization (WHO) objective of both parents’ HIV prenatal screening [1].

At a time when pertussis and measles are spreading across Europe and causing serious infections in newborns [18] [19], two-thirds of participants were overdue for pertussis vaccination, which is more than what was previously reported in France [20]. More than half of overdue participants received vaccination updates on site. The study’s vaccination allocation was insufficient to cover all needs, leading to preference being given to men who would have to pay for their vaccines at the pharmacy. Although we cannot measure the impact of vaccines prescribed without administration, the final coverage rate would probably have been higher if we had had the means to vaccinate all willing fathers immediately on-site.

We have shown elsewhere that immigrants had the highest rate of consultation acceptance, especially the most precarious ones [16]. This study shows that, for each indicator, the effects of the consultation were more frequent among socially disadvantaged immigrant participants. Socially disadvantaged immigrants therefore cumulate greater attendance and more health effects in the event of attendance. Whatever the outcome considered, the absence of any health coverage was always most strongly associated with its occurrence.

Immigrants, especially those born in Sub-saharan Africa and those in France for less than 7 years, were overrepresented among participants with any unknown or lost to follow-up pathology. Some factors partly explain this overrepresentation: tropical parasites screening was restricted to participants who had grown up in Sub-saharan Africa; some infectious diseases, such as hepatitis B, are far more prevalent in endemic regions, but uncovered health needs were probably also more prevalent in this population. During the first seven years of residence in France, most immigrant men face hardship, while the proportion of settled immigrants grows after this period [21].

After adjustment on region of birth, school level, age and employment, having no health insurance coverage remained associated with both identification of medical diagnoses and referral to healthcare professionals, while having State Medical Assistance no longer was. Prior State Medical Assistance acquisition often reflected previous contact with the healthcare system. Compared to undocumented persons with no health insurance coverage, those with State Medical Assistance also have easier access to the healthcare system. Therefore, State Medical Assistance is both a tracer and a factor in access to care. Our results suggest that State Medical Assistance leads to earlier diagnosis and better retention in care. In Germany and in the Netherlands, the absence of health insurance coverage in undocumented immigrants was consistently associated with late presentation of chronic diseases and discontinuity of care [22] [23].

On-site vaccination was more frequent among immigrants than among French-born participants, more immigrants being overdue and more immigrants having no opportunity to get a vaccine update elsewhere, for lack of full health insurance coverage or of effective access to other healthcare professionals.

While the need to improve men’s health outcomes is well established [24] [25], PARTAGE is, to our knowledge, the first structured health intervention built to address adult men’s health, by using the symbolic event of expecting a child. However, our study had some limitations: we demonstrated the feasibility [16] and the positive effects of paternal prenatal consultation, but not its replicability, as our proactive approach is not directly transferable to routine care.

No similar experience has been reported. Published interventions focus either on involving future fathers in prenatal care [26] or in parenthood [27], or on testing them for HIV [28] [29] [30] [31]. A paternal prenatal screening for HIV and STIs was once experienced in Europe, but the attention paid to fathers was restricted to the risk of infectious disease transmission. Men attending prenatal ultrasound in London with their pregnant partner were offered on-site serology for HIV, syphilis, hepatitis B and hepatitis C and urine testing for *Neisseria gonorrhoea* and *Chlamydia trachomatis*. The proportion of men who had never been tested for HIV before was higher than ours (59%), and the acceptance rate lower (35%); as in PARTAGE, no HIV infection was discovered, but other STIs were detected and managed. The authors concluded hence, that the initiative was useful [32]. Despite a plea for better vaccination coverage of both parents against pertussis [18], interventions aimed at vaccinating future fathers remain scarce [5].

Reaching future fathers and promoting a consultation they do not expect to be offered is a challenge, especially without the support of a dedicated team as in PARTAGE. A new phase of the program, PARTAGE 2, is currently being launched in Montreuil city to assess the conditions for real-life deployment. Another unanswered question is how to identify future fathers early in pregnancy without disempowering women. For policy makers, scaling-up would imply rewarding this complex consultation in proportion to the medical time it requires.

## Conclusion

Paternal prenatal consultations were useful to the whole study population, by creating a systematic and well-accepted opportunity for biological screening and vaccination catch-up. It also led to identification of undetected illnesses’ and to resumption of care of patients with chronic diseases lost to follow-up, and to linkage into healthcare. Health benefits were greater in the most socially disadvantaged future fathers, who had more health needs. These were also enabled to meet social workers when necessary. Therefore, paternal prenatal consultation could help reduce both gender and social inequalities in health.

Scaling-up this consultation requires creating adapted tools to reach future fathers, taking into account the time constraints faced by men and building a free-of-charge package including a long and complex medical consultation, biological tests and vaccines.

## Funding

The PARTAGE study was financed by the French Agency for research on AIDS, viral hepatitis, tuberculosis and emerging infectious diseases (ANRS-MIE) (research team’s salaries) and by the French Society against AIDS (SFLS - implementation and evaluation of a Saturday consultation outside hospital walls). Montreuil’s hospital also received a donation from Gilead Sciences, Inc. to develop PARTAGE’s communication and IT tools.

## Aknowledgments

The authors thank all participants.

The PARTAGE working group includes, in addition to the authors: Clotilde Trevisson, Anne-Laurence Doho, Patricia Obergfell, Djamila Gherbi, Emilie Daumergue, Naima Osmani, Sandrine Dekens, Oumar Sissoko (ARCAT), Virginie Supervie, France Lert, Bruno Renevier, Thomas Phuong, Stéphanie Demarest, Ngone Diop.

Thanks to Christophe Michon, Nathalie Lydié, Joanna Orne-Gliemann, Laurent Mandelbrot, Corinne Taeron, Nicolas Derche, Gwenaelle Morvan, Bernadette Rwegera, Mélanie Jaunay, Ruth Foundje Notemi, Coraline Delebarre, Elisa Wardzala, Caroline Regnier, Clelia Fouache, Hiba Boufares, Paul Chalvin, Perrine Bonnefoy, Priscillia Ribouchon, Abdelkrim Imechket, Pauline Aubry, Francis Bouvier, David Benhammou, Véronique Doré and Yoann Allier (ANRS-MIE), Frédéric Goyet (ARS Ile de France), the city of Montreuil, Marie Pastor (Conseil Départemental de Seine Saint-Denis), Marie Némon, Pascal Biehler, Nadia Ben Lagha, Justine Bellinger, Isabelle Auperin, Guy Nielsen.

## Conflict of interest

The authors have no competing interests.

## Key points

● In disadvantaged European areas such as ours, social deprivation and gender norms contribute to keeping men out of the healthcare system: targeting fathers during pregnancy is an opportunity to address both social and gender inequalities in health.
● Eligible participants largely accepted HIV testing (99%).
● In a context of European outbreaks, 72% of future fathers were out-of-date for measles or pertussis vaccination: 60% of this delay was addressed on site.
● This offer, which led to the identification of a new diagnosis for 18% of participants and a referral to a healthcare provider for 17%, mostly benefited disadvantaged immigrants.

## Patient and Public Involvement section

A pilot study was conducted from 2017 to 2020, limited to HIV and STI screening in Montreuil hospital prenatal care waiting rooms: future fathers’ extra needs were collected by the pilot research team and used to build PARTAGE intervention;

An association providing support to immigrant HIV-infected women was involved in drafting and validating the maternal and paternal research questionnaires;

The questionnaires were tested with five pregnant women and five future fathers before starting the study;

All participants were asked if the prenatal consultation offered met their needs and what could be done to get as many men as possible to accept it (see Paternal Questionnaire, Supplementary file 3).

After PARTAGE, several focus groups were held with future fathers from Montreuil city, to co-construct the consultation scaling-up at municipal level (PARTAGE 2 study).

## Authors’ contribution

Conceptualization: Pauline Penot, Annabel Desgrées du Loû, Gaëlle Jacob, Audrey Guerizec Funding Acquisition: Pauline Penot, Pierre-Etienne Manuellan

Methodology: Pauline Penot, Annabel Desgrées du Loû

Formal analysis: Pauline Penot, Annabel Desgrées du Loû, Yazdan Yazdanpanah, Raya Harich

Investigation: Pauline Penot, Valérie-Anne Letembet, Gaëlle Jacob, Audrey Guerizec, Anne Simon, Miguel de Sousa Mendes, Raya Harich

Project administration: Pauline Penot, Gaëlle Jacob, Audrey Guerizec, Pierre-Etienne Manuellan, Bruno Renevier, Raya Harich, Annabel Desgrées du Loû

Writing-original draft: Pauline Penot, Annabel Desgrées du Loû, Yazdan Yazdanpanah, Miguel de Sousa Mendes

All authors reviewed the current manuscript, gave final approval to it submission and agreed to be accountable for all aspects of the work.

## Data Availability

All data produced in the present study are available upon reasonable request to the authors

## Notes

### Competing Interest Statement

The authors have declared no competing interest.

### Clinical Trial

NCT05085717

### Author Declarations

The French Data Protection Authority (CNIL, registration number 921135) and the French Personal Data Protection Committee (Comite de Protection des Personnes, registration number 21.01.19.44753) gave ethical approval for this work.

