## Supplementary File 1 for "Does a prenatal consultation dedicated to fathers’ health widen men’s access to prevention and care? A monocentric interventional research in the Paris metropolitan area"

### PARTAGE Project

**Title : PARTAGE (“Prevention, Accès aux droits, Rattrapage vaccinal, Traitement des Affections pendant la Grossesse et our l’Enfant”). Acceptability and determinants of acceptance of a male prenatal consultation, including screening, vaccination catch-up and access to health insurance coverage, offered to future fathers at the Montreuil hospital, Seine-Saint-Denis.**

#### 1. Research objectives and innovative character :

Insufficient screening and delayed diagnosis play a major role in maintaining the HIV epidemic in France. Although the first decline in incidence has been observed in Paris among men who have sex with men (MSM) born in France, no significant decline has been recorded among heterosexuals, whether born in France or abroad (Santé Publique France 2019): major gender inequalities combined to social inequalities make foreign-born heterosexual men the population diagnosed with HIV the latest.

The effectiveness of antenatal HIV screening, offered to women for every pregnancy and widely accepted, plays a large role in this gender gap: 18% of women newly diagnosed in France in 2017 discovered their HIV with a pregnancy test (156 women out of the 860 for whom the reason for screening was given on the declaration to the French Public Health Agency). Expecting a child could be an opportunity for the couple to undergo joint screening. However, although a consultation and biological check-up for fathers-to-be is supposed to be possible and fully covered by the French health insurance system, men's health is not taken into account in standard French prenatal care.

We propose an interventional research to implement a prenatal consultation for all future fathers at the Montreuil hospital in Seine Saint Denis, France. We will study the feasibility and processes of this implementation.

Our tertiary care maternity unit attracts a population marked by immigration, often living in precarious conditions. The pilot phase of this project revealed a wide range of needs in terms of prevention and access to healthcare among future fathers. We will therefore assess the impact of this male consultation on HIV screening, but also on screening for other infections (viral hepatitis, sexually transmitted bacteria), vaccination coverage, health insurance coverage and access to a general practitioner for subsequent follow up.

The aim of this intervention is threefold: to reduce the time between HIV infection and diagnosis in men, to improve or preserve their general health, and to reduce the transmission of infectious diseases to the mothers and children. The ultimate aim is to transfer this male prenatal consultation into current clinical practice, as it is theoretically provided for by the health insurance scheme but not yet organized nor implemented.

This research is innovative as, to our knowledge, no team has so far studied the impact of offering a dedicated prenatal consultation to all future fathers.

#### 2. Situation of the subject in the national and international context, and involvement of teams in the subject

All over the world, healthy men have less contact than women with the healthcare system (Greig, Kimmel, and Lang 2000). Self-identified heterosexual men are diagnosed with HIV later than women (Kigozi et al. 2009). (Bonjour et al. 2008) (Op de Coul et al. 2016). Interventions to promote screening during their partner's pregnancy have mainly been tested in sub-Saharan Africa and are summarized below.

It is important to point out that the risk of acquiring HIV increases in pregnant women: this risk doubles during the first trimester and triples at the end of pregnancy (Thomson et al. 2018) . A primoinfection in pregnancy carries a high risk of maternal-fetal transmission: it often escapes the serology performed at early stages of pregnancy, and the high viral load increases the probability of infection in the newborn baby (Wertz et al. 2011).

##### 2.1. Sub-Saharan Africa :

The effectiveness of couple-oriented prenatal HIV screening in reducing maternal-fetal transmission has been shown in countries with high HIV prevalence (Dunlap et al. 2014) (Aluisio et al. 2016) (Crepaz et al. 2015). However, couple adherence to a joint screening approach remains low, below 20% in most interventions carried out in different sub-Saharan African contexts: 16% in Tanzania (Becker et al. 2010)15% in Kenya (Farquhar et al. 2004)9% in Uganda (Semrau et al. 2005)16% in Ethiopia (Haile and Brhan 2014).

Fear of violence or repudiation in case of positive HIV testing in the presence of their husbands was often cited by women as a reason for their reluctance to bring their husbands in: yet studies have shown that couple-oriented prenatal screening does not increase violence against women, and even reduces the risk of it, compared with individual screening of pregnant women (Mohlala, Boily, and Gregson 2011) (Semrau et al. 2005) (Orne-Gliemann et al. 2013).

Women didn't always feel legitimate in relaying a screening offer, because wives don't talk about sexual matters and because the proposal could be seen as an injunction to their husbands (Falnes et al. 2011) (Larsson et al. 2010). In Cameroon, as part of the ANRS 12127 PRENAHTEST trial, the counselling given to pregnant women upon receipt of their HIV serology results has been strengthened. The aim was to enable them to define concrete strategies for telling their partner about the result and inviting him to take part in screening: this intervention increased the rate of men getting screened from 14% (control arm) to 25% (Orne-Gliemann et al., 2004). (Orne-Gliemann et al. 2013). In a Kenyan survey, participating women preferred their husbands to be directly invited by the healthcare structure (Osoti et al. 2015). In Uganda, an invitation letter written to husbands increased couple prenatal visits by 10%, and almost all men who attended accepted joint HIV testing (95%) (Byamugisha et al. 2011). In South Africa, 32% of future fathers to whom an invitation letter had been sent accepted joint testing (Mohlala et al. 2011).

Even so, men have difficulty finding their place in health facilities dedicated to women and children: care facilities are identified as women's spaces, and care providers are not always welcoming to men. What's more, the opening hours of antenatal clinics and the long waits imposed on users make them difficult to access for men, who are supposed to provide for the family's financial needs (Ditekemena et al. 2012) (Nkuoh et al. 2010). Noting that gender norms and professional obligations kept men away from prenatal health centers, some teams have offered to test them at home (Osoti et al. 2015), in bars (Ditekemena et al. 2012) and even in churches, where the success of the intervention was spectacular: in southern Nigeria, celebrations organized by the clergy ("Baby Shower") resulted in 86% of future fathers approached being screened, a rate twelve times higher than the control arm.

Meanwhile, while the entire burden of pregnancy and child health falls on women (Haile and Brhan 2014) (Nkuoh et al. 2010), women's autonomy, and in particular their freedom to honor their consultations, remains controlled: recourse to an institution (the public sphere) is a matter for male decision-making power (Baiden et al. 2005) (Kiarie et al. 2003) (Homsy et al. 2007).

.

#### 2.2. In Europe

The paucity of literature and organized initiatives on prenatal screening extended to men in Europe and the United States is probably linked to the low incidences of HIV in these parts of the world. Yet in Europe, including the European Union, the heterosexual epidemic is only slowly receding, and immigrants, both men and women, are numerous among those newly diagnosed.

International recommendations have long called for prenatal screening to be extended to couples. As early as 2010, the World Health Organization (WHO) affirmed that "male partners play an equally important role in the scale-up of PMTCT [prevention of mother-to-child transmission] services" (World Health Organization 2010). The same year, President's Emergency Plan for AIDS Relief (PEPFAR) experts recommended increasing male participation in PMTCT to improve its effectiveness (PEPFAR 2010).

Based on these findings and recommendations, a London team offered screening to all men accompanying their pregnant partners to obstetric ultrasounds for six months in 2011. 35% of the men approached accepted, representing 18% of the 2,400 pregnancies monitored by ultrasound over the period. Half of the men tested had no previous HIV serology; no HIV infection was diagnosed, but several hepatitis B, C and urethral carriages of *Chlamydia trachomatis* were discovered (Dhairyawan et al. 2012).

In northern Sweden, in-depth interviews were carried out in 2009-2010 with twenty partners of pregnant women, in order to identify the obstacles to extending prenatal screening to future fathers. Among them, twelve had accepted screening (no HIV found, one urethral carriage of *Chlamydia trachomatis*), eight had refused. They perceived themselves as being at low risk of HIV and were unaware that they could affect the pregnancy or the health of the newborn by transmitting a silent infection to their partner. At the end of the interviews, 18 of the 20 said they were in favor of systematic screening for future fathers, and most pointed to the midwife as the most legitimate professional to convince future fathers of the need to be screened (Christianson, Boman, and Essén 2017).

#### 2.3. National and local context

*In France*

PrEP (*Pre-exposure prophylaxis*), TasP (*Treatment as prevention*) and the increased availability of screening (creation in 2016 of free information, screening and diagnosis centers - CEGIDD) primarily benefit people who self-perceive themselves to be at risk of HIV and who have access to prevention facilities: the number of new HIV diagnoses fell by 28% among MSM born in France and residing in Paris between 2015 and 2018, but no significant decline was observed among people born abroad, particularly heterosexuals, over the period.

Joint screening of future parents during each pregnancy is recommended by the French National Authority for Health ("Haute Autorité de Santé", HAS) (HAS 2009). A prenatal consultation dedicated to future fathers, including biological tests, is supposed to be free of charge. Unfortunately, this recommendation is hardly applied at present: in 2018, in Seine-Saint-Denis, only 0.2% of men identified as future fathers by the local level of the health coverage system (CPAM) benefited from a consultation referenced as "prenatal" (data from

Julien Bordron, CPAM du 93, who considered that practitioners' lack of familiarity with the dedicated nomenclature had probably biased the numerical estimate).

In the 1990s, Professor Mandelbrot tried to set up a consultation dedicated to future fathers at the Port Royal maternity hospital in Paris. His team soon gave up, noting that the men most at risk of HIV infection were not using it. The experiment was not published.

What's more, maternal serology at the end of pregnancy is virtually never prescribed in France, where women's sexuality during pregnancy is rarely questioned in consultation: the best way to limit pregnancy-related transmissions would therefore be to stop "ignoring half of the pregnancy equation" (Mohlala et al. 2011).

*In Seine-Saint-Denis*

Seine-Saint-Denis is the poorest department in mainland France. Its demography is strongly marked by immigration, and particularly recent immigration: 36% of women and 39% of men aged 15 to 54 who live there were born abroad (INSEE 2019). In 2013, 102,000 people born in sub-Saharan Africa were registred in the department.

It is the second department most affected by HIV epidemic in mainland France: 9,000 HIV-infected people live in the region, and around 600 discover their seropositivity each year. This represents an incidence rate at least 3 times higher than the French average. The epidemic is 74% heterosexual (France Lert 2017). The prevalence of sero-ignorance among adult men born in sub-Saharan Africa living in Seine-Saint-Denis has been estimated at 1% (Marty et al. 2018).

In 2018, 4008 babies were born at the Montreuil tertiary care maternity unit, corresponding to some 3840 deliveries: 21 involved HIV-positive women who were monitored jointly by infectiologists, obstetricians and pediatricians during their pregnancy and postpartum.

A pilot project has been working since October 2018 to build a prenatal HIV screening intervention for fathers of unborn children at the Montreuil hospital center.

All male partners of women being monitored for an active pregnancy were offered screening, either directly (information at antenatal clinics and visits by the research team to waiting rooms), or through their partner (oral and written information on the benefits of screening, and access to the CEGIDD). Screening could be carried out by rapid diagnostic test (TROD) or serology, either immediately in a dedicated room in the maternity ward, or during opening hours at the CeGIDD (free information, screening and diagnosis center for human immunodeficiency virus infections, viral hepatitis and sexually transmitted infections).

Between October 2018 and August 2019, 420 men were screened for HIV (by rapid HIV test [TROD] or serology) and, for some, for viral hepatitis and bacterial sexually transmitted infections (STIs). A quarter of them were born in sub-Saharan Africa, 19% in North Africa and 31% in mainland France (see appendix 1, graph 1). The 56 nationalities represented at birth reflected the extraordinary diversity of origins of young adults living in Seine-Saint Denis. The median age of the men included was 33 (interquartile range [28-38]. Just over half (51%) were expecting their first child. Nearly a third (32%) met at least one of the following three criteria of extreme precariousness: no formal income-generating activity, no right of residence or no social protection (Appendix 1, graph 2).

Nearly half (45%) had never had any HIV test. This proportion, which was initially higher, gradually fell as the program was implemented, in parallel with the drop in the proportion of highly precarious new arrivals received. The first fathers reached were more remote from the prevention and screening offer than subsequent ones: the generalization and repetition of the proposals led better-integrated fathers to accept prenatal screening.

The main ways of finding out about the service were by visiting the three antenatal clinic waiting rooms (45%), and direct referral by the obstetrician-gynecologist or midwife during an antenatal consultation or obstetric ultrasound (44% of referrals), while the information given to pregnant women about the need and procedures for screening their partners led to few men being screened (only 8% of fathers screened came without having been directly solicited) (Appendix 1, graph 3). However, the rate of refusals on the street was high, and obstetricians and midwives were themselves difficult to mobilize. Maternity wards are still a feminine space: the men present were not very visible, and the nurses in charge of prenatal care were not accustomed to approaching them: in a way, the men were outside their usual sphere of responsibility, outside their field of competence.

The fathers actually screened all expressed great satisfaction with the initiative, and most spontaneously welcomed the opportunity to participate in pregnancy monitoring. Broader needs in primary and secondary prevention of infectious diseases emerged: insufficient vaccination coverage, lack of contact with a health professional since arriving in France for many immigrants, need to meet with a psychologist or a social worker.

This initial analysis led us to develop a new, systematic offer of preventive consultations dedicated to future fathers, in order to preserve or improve their health and to increase men's acceptance of HIV testing; it convinced us of the importance of facilitating access to this consultation for future fathers, in particular by locating it outside the maternity unit, or even outside the hospital itself, and by scheduling it at times compatible with a professional activity; the pilot also highlighted the limits of a prevention offer relayed by pregnant women, and the need to address men and couples directly, provided that the women had given their consent. Finally, it showed us that the most legitimate people to deliver this message were the physicians and midwives in charge of the pregnancy.

#### 3. Working hypotheses and specific objectives

The differential acceptance of HIV serology between pregnant women and their partners is linked to a gender stereotype: the health of the unborn child is the sole responsibility of the pregnant woman. This stereotype is reinforced by the way prenatal care is organized.

A systematic offer of screening, based on shared parental responsibility rather than sexual risk-taking, would make it possible to reach asymptomatic heterosexual men who perceive themselves as being at low risk of HIV: during pregnancy, HIV can be "normalized", brought back to the status of other infections systematically screened in pregnant women, which share with it not their mode of acquisition (respiratory for rubella, telluric and alimentary for toxoplasmosis), but a common risk to infect the fetus.

When the caregiver in charge of prenatal follow-up is convinced, acceptance of screening is high: doctors and midwives must therefore be trained to take an interest in the partner's health. When a medical chart without the father’s serologies registred will be considered incomplete, couples' habits will change.

Men seen in individual prenatal consultations should have a high level of acceptance of the examinations prescribed to them. Similarly, if a chronic infection is discovered, our hypothesis, inspired by the HIV cascade in France, is a high retention in the healthcare system.

This male prenatal consultation exists but is not organized: it is possible to implement it, provided that the constraints placed on men are taken into account.

Thus, the present research aims to:

- Establish the level and determinants of acceptance of this male prenatal consultation, through different modalities (in the maternity unit, elsewhere in the hospital, in the heart of town; during the day, in the evening or on Saturdays; with or without an appointment).

- Describe the socio-demographic characteristics of fathers attending antenatal clinics, compared to those who did not attend (using the maternal questionnaire); describe the reasons given by men who did not attend, when contacted.

- Compare the answers given by women at the time of their enrolment in the maternity unit (history of HIV testing of their partner, possible results, expected acceptability of the male prenatal consultation, expected acceptability of a serological test in the context of their future paternity) with the reactions and answers given by men (subscription to the offer of consultation, previous tests and possible results, acceptance of a prenatal biological test).

- Study the proportion of future fathers who :

    - accept HIV testing during this prenatal consultation (by analyzing the situation with regard to previous testing: never tested/ tested at least once/ tested several times) ;
    - are seeing a physician for the first time since arriving in France;
    - do not have an attending physician;
    - open health insurance coverage rights thanks to this prenatal consultation;
    - require medical intervention following the consultation and, of these, how many are cared for (treatment or follow-up);
    - don't have their vaccinations up to date, and how many of them have been vaccinated?
    -

#### 4. Project presentation

##### A. Study design

We propose a monocentric cross-sectional study without control arm, at the André Grégoire intercommunal hospital in Montreuil (Groupement Hospitalier de Territoire [GHT] Grand-Paris Nord Est) for 15 months, followed by adaptation and implementation of the intervention in daily clinical practice.

##### B. Study population

All men living in the Ile-de-France region whose partner is in care at Montreuil maternity unit for a progressive pregnancy are eligible for a prenatal consultation. Minors are excluded from the study but benefit from the intervention.

#### C. Intervention

The meeting with the PARTAGE project interviewers will be integrated into the women's maternity trajectory. The interviewers will present the project to eligible women and offer them to respond to a face-to-face questionnaire that will include their socio-demographic and marital characteristics (maternal questionnaire). Women who consented will be asked to provide the future father's contact details. The project will be presented directly to the couple when the future father is present, and the consultation appointment will be possible to schedule immediately.

When a woman has no time to answer the maternal questionnaire the same say, a telephone appointment will be offered, or the woman will be met at her next prenatal consultation, unless she has given birth in the meantime: childbirth censors the women's offer to take part in the research.

An information letter will be given to all eligible pregnant women. An explanatory e-mail will be sent to men for whom pregnant women have provided an e-mail address, with an online appointment platform link, a telephone number available 6 days a week and a dedicated e-mail address to schedule the prenatal prevention consultation. It will be mentioned in both letter and e-mail that if no spontaneous appointment is made, the father will be called the following week by the research team. One week later, or on the Monday following inclusion when the women did not provide an e-mail address, fathers will be called by a research midwife and offered an appointment either during the day or in the evening, either on weekdays or Saturdays, either at the hospital or at the health center located in the city center, easily accessible by public transport. They will be asked to bring their health or vaccination booklet, if they have one, and any biological tests they may have.

Fathers will receive a text reminder before their appointment. If they fail to attend, they will systematically be called again and given the option of either refusing the consultation (their reason for refusal will be collected and their contact details will be deleted), or rescheduling and being called again in case of another missed appointment.

Fathers will be seen by a doctor or a midwife, trained in the project and not involved in the partner's prenatal care. A research questionnaire will be included in their consultation chart (paternal questionnaire). Blood pressure will be measured; clinical examination will be carried out in case of symptoms. The biological check-up will systematically include a HIV serology, unless a test was carried out during the current pregnancy, and others analysis be adapted to individual's exposure and medical history. Vaccines will be updated and subsequent doses will be scheduled when required; fathers lacking health insurance coverage will be offered to meet a social worker, in order to obtain it. Test results will be delivered either face to face, by telephone or by e-mail, according to men’s choice. Depending on their needs, men could be referred to a health mediator, a psychologist or any hospital specialist. If they had no previous medical follow up, they will be referred to a general practitioner.

The strictly confidential nature of all information given to the team and of anything that may be discovered in relation to their state of health will be explained to both future parents: in particular, no biological results will be divulged to the partner without the patient’s consent.

Pregnant women, future fathers accepting and refusing the offer and healthcare professionals working in the maternity unit will be offered to meet the sociologists in charge of the qualitative part of the project, for semi-structured, non-nominative individual interviews lasting around an hour. The sociologists will also be involved in participatory observation at various levels of the project: in maternity unit waiting rooms, with interviewers, in paternal prenatal consultations.

**E- Judgment criteria**

Primary endpoint: Proportion of future fathers who accepted the prenatal consultation among those eligible. Men involved in the pregnancy, living in the Ile-de-France region and whose pregnant partner has provided accurate contact details - telephone or e-mail – for, or who have themselves been approached during the couple's visit to the maternity unit, will be considered as eligible.

Secondary endpoints:

Number of future fathers who accept a biological test including HIV serology, compared to the number of fathers eligible, over 15 months; proportion for whom HIV seropositivity, hepatitis B or C, an STI or another infection will be discovered and, among them, proportion retained in care (HIV, HBV, other chronic infection) or cured (syphilis, hepatitis C, chlamydia, gonococcus, other acute or curable infection) at 12 months;

Other infectious disease prevention interventions: proportion of updated vaccination schedule (required/effective), opening of health insurance coverage (required/effective), referral to another professional; proportion of future fathers declaring an attending physician at the start and at the end of the intervention.

There is no judgment criterion related to HIV infection, as the expected number of HIV discoveries is low. The only estimate of undiagnosed HIV epidemics we have for our territory concerns men from sub-Saharan Africa: 25% of the men included in the first phase of the pilot came from this region of the globe, i.e. a projection of 3 HIV discoveries in this population for every 1000 inclusions (of which 25% were men born in sub-Saharan Africa).

**F - Data collection, statistical analysis, ethical considerations**

*Data collection*

Pregnant women and their partners will receive oral and written information about the project and their non-opposition will be collected.
Maternal questionnaire will be recorded into ORBIS© software (Agfa Healthcare, Bonn, Germany), the hospital's medical data system.
Future fathers designated by pregnant women will be registered: they can choose at any time to be deleted, or to be designated by a pseudonym of their choice (but warned that the administration of a vaccine or treatment require a consolidated identity). A pregnancy number will link them to their female partner. Paternal questionnaire will also be recorded into ORBIS© software.
Data collected by the health anthropologist will be processed separately, as the ORBIS© software is only accessible to health professionals.
This collection method has been validated by the CNIL and the Comité de Protection des Personnes Nord-Ouest II (National number: 2020-A03198-31).

*Statistical aspects*

Out of 200 randomly selected obstetrical files in 2018, 192 women declared a father involved in their pregnancy and living in the Ile de France region. This estimate, projected over the 5000 deliveries in fifteen months, brings the number of fathers theoretically accessible to prenatal intervention to 4800. Taking into account the fathers who will not receive the information, those who will refuse the consultation, those who will accept the consultation but not participation in the research and those who will choose to consult their general practitioner or will have already seen him by the time the intervention is proposed to them, we have set ourselves the target of including at least 20% of accessible partners, i.e. at least 960 future fathers in 15 months.

We will compare the characteristics of the future fathers included with those of the non-included ones (maternal questionnaire).

We will establish the following cascade: number of women attending antenatal clinics at Montreuil hospital; number of women giving details of a partner involved in the pregnancy and living in the Ile-de-France region; number of women included in the research (having completed questionnaire 1); number of partners contacted for a proposal for antenatal consultation; number of spouses actually seen for prenatal consultations; number of spouses screened for HIV and possible co-infections; number of HIV and hepatitis B diagnoses made; number of fathers medically monitored six months after diagnosis of chronic infection.

Statistical analyses will be carried out using STATA© 12.2 software (StataCorp, College Station, Texas).

*Ethical considerations*:

Particular attention will be paid to reciprocal confidentiality: HIV-positive women who have been diagnosed during previous pregnancies and who have maintained confidentiality may be confronted with the discovery of their own seropositivity by their partner. The male prenatal consultation has thus to be understood as a systematic offer, integrated into the normal monitoring of all pregnancies. The widespread promotion of the project by posters in the maternity unit should protect HIV women from the suspicion of an intervention targeted at their partner.

Similarly, a man who knows or suspects he is HIV-positive may fear disclosure of his status. For HIV-positive men already diagnosed: a viral load will be taken during the prenatal consultation, the vaccination schedule will be updated and co-infections checked if necessary. If a man discovers that he is HIV-positive, or if his viral load is detectable, he will be seen by the infectiologist and encouraged to share the information with his partner, with a proposal for medical and associative support for this process. Antiretroviral treatment will be initiated or re-evaluated, and compliance reinforced if necessary. If a man with a detectable viral load chooses to remain secret, the obstetrical team will be informed that his HIV status is unknown: in this case, the French recommendation will be applied, and his partner will be offered a new HIV serology test in the third trimester (HAS 2009). During the pilot phase, six men had positive or doubtful ELISA HIV serologies, all of which were subsequently invalidated by Western blot, viral load or analyses carried out at the associated Rouen national reference center: all 6 immediately informed their partner of their uncertain result.

Men who are carriers of *Chlamydia trachomatis* will be given a single course of treatment directly, and oral and written information will be provided on: the frequency of chronic, asymptomatic carriage, which may indicate a very long-standing infection; the risk of transmission to mother and child; the indication that the same treatment should be given to pregnant women, without risk to the pregnancy; the proposal to communicate the prescription directly to the midwife or doctor in charge of prenatal follow-up, if the future father agrees. At the same time, we are advocating with our gynecologist-obstetrician colleagues that pregnant women under the age of 26 should be systematically screened for vaginal carriage, in line with national recommendations (HAS 2018). A gonococcal carriage will be treated in a similar way, with the difference that screening may be offered to the pregnant woman before treatment if the last sexual intercourse is more than seven days old, due to the discomfort of an injectable treatment.

If hepatitis B, C or syphilis is discovered, the patient will receive medical care, oral and written information on the risks of transmission and the steps to be taken by the partner, and a proposal to accompany disclosure. For hepatitis B: verification (and possible extension to the three markers) of maternal serology is sufficient to assess the risk of transmission to the unborn child. If the mother is negative for all three markers, and if the father consents to his status being communicated to his partner, the pregnant woman will be offered immediate vaccination.

This research protocol has been approved by the Comité de Protection des Personnes (CPP) Nord-Ouest II (registration number: 21.01.19.44573, approval for substantial modification: 21.05022944753). Protocol is registred on http://clinicaltrials.gov (NCT05085717).

#### 5. Involvement of each team in research

**Hospital team**

The project will be run by two teams at the Montreuil hospital: the CeGIDD on the one hand, and the maternity unit, headed by Dr Bruno Renevier, on the other.

Doctor Pauline Penot, an internist in charge of the research, will divide her time between the CeGIDD, infectious disease consultations and 0.2 full-time equivalent (FTE) dedicated to the project. Gaëlle Jacob, midwife at the maternity unit and at the CeGIDD, will be on the project on a 0.5 FTE basis (for 15 months) then 0.4 FTE basis (for 12 months). They will share project coordination and the following tasks:

For 15 months: scheduled and unscheduled male prenatal consultations, coordination of the research team, monitoring of inclusions, follow-up on contacts made with eligible fathers, control of data collection, follow-up of legal approvals, follow-up of biological results and their reporting, follow-up of referrals and vaccinations.

During the following 12 months: data processing, oral and written scientific communication of results, implementation of male consultation in routine clinical practice.

Audrey Guerizec, a midwife specialized in public health, will be involved in the research on a 0.6 FTE basis to: carry out male prenatal consultations, supervise the legal procedures linked to the project, take part in the animation and cohesion of the research team, the articulation of the different stakeholders and the planning of consultations (for 15 months); follow the care pathways of men diagnosed with a chronic infection during the project, check that they obtain health insurance coverage; take part in data processing, its valorization and the transfer of the intervention to real life (for 12 months).

Research midwives will be required to collect biological samples as part of the project, and may at any time refer to a doctor an expectant father who is symptomatic, or whose test result comes back positive or abnormal.

Stéphanie Demarest, an affective life counselor, works full-time in the hospital's family planning unit (financed by the department) and has been trained in HIV prevention and screening through a partnership with CEGIDD. She will contribute 0.2 FTE to the project, coordinating calls and e-mails sent to future fathers, scheduling their consultations, checking that they have attended and contacting them again if they have not.

The CEGIDD's nurses and social worker are funded by the Agence Régionale de Santé d'Ile de France (ARS-IDF), which pilots the healthcare system at the regional level and funds every CeGIDD. Nurses will collect biological samples when consultations are carried out at the CeGIDD. The social worker will receive all fathers identified by the consultants as needing support to open up social rights. At the healthcare center, samples will be taken by nurses employed by the city of Montreuil, under a partnership agreement.

One or two doctors with experience in sexual health will be recruited in the first year to provide male prenatal consultations on and off hospital premises.

Two full-time interviewers will be recruited over a 15-month period to administer maternal questionnaires. Two sociologists will explore the obstacles, facilitators and determinants of pregnant women, fathers and staff adherence to the project. In the second year, a statistician will support the research team in processing and valorizing the results.

**Research support - associate experts**

Dr Pauline Penot is an associate researcher at the Population and Development Center (CEPED, multidisciplinary research unit), as part of the Vulnerabilities and Gender Relations team, headed by Annabel Desgrées du Loû, who will provide her with support. Dr. Christophe Michon, infectiologist is a public health physician working at the French National Health Direction (DGS). Prof. Laurent Mandelbrot is the head of the gynecology-obstetrics department at Colombes hospital. They will be invited to join the scientific advisory board, which already includes: Nathalie Lydié (Santé Publique France), Joanna Orne-Gliemann (Institut de Santé Publique, d'Epidémiologie et de Développement, Bordeaux), France Lert (CEPED), Virginie Supervie (Inserm).

**Partnerships and funding**

This project will be carried out in close collaboration with the institutions in charge of prevention in our region: the ARS, and in particular its Seine-Saint Denis Territorial Department, which funds CeGIDD staff and biological tests, the Department Council (CD 93), which funds hospital-based Family Planning staff, and Santé Publique France.
Dr. Manuellan, Director of Health for the City of Montreuil, is providing the project with a consultation area within the city’s healthcare center, located in the same building as the city's administrative services.
The GHT Grand-Paris Nord-Est is supporting the project through its clinical research unit, which will be able to receive research funding, the Montreuil site's communications officer (Sophie Villattes), IT support (Vincent Bruneau) and the participation in the project of several hospital practitioners, including Dr Raya Harich, from the biology laboratory unit.

PARTAGE project was funded by:

- The French National Agency for Research on Aids, viral hepatitis, tuberculosis and emerging diseases (ANRS-MIE)
- The French Society Against AIDS (SFLS)
- The Ile de France Health Agency (ARS-IDF)
- The Seine Saint Denis Department Council (CD 93)

It received support from: Montreuil Hospital (GHT Grand Paris Nord Est), the city of Montreuil, the associations IKAMBERE and ARCAT, the GILEAD Foundation.

#### 6. Expected impact

The ultimate aim of this research is to move from theory to practice. This real-life transfer meets an urgent need, in our territorial context, for access to men who are far removed from prevention and care services, particularly immigrant men from sub-Saharan Africa.

Taking men's health into account during pregnancy will help to reduce the undiagnosed epidemic of HIV and viral hepatitis, reduce pregnant women's exposure to HIV, address gender inequalities by distributing responsibilities more equitably between the two parents, avoid vaccine-preventable infectious diseases and facilitate the integration of men in the healthcare system.

To achieve these public health benefits, a paradigm shift is needed. Our mobilization aims to ensure that future parents include a consultation dedicated to the father in their representation of prenatal care, but also that prenatal caregivers ensure that the future father has been seen and screened during the current pregnancy. In this way, we hope to improve men's health, preserve women and children’s safety and reduce social and gender inequalities in health.

Falnes, Eli Fjeld, Karen Marie Moland, Thorkild Tylleskär, Marina Manuela de Paoli, Sia E. Msuya, and Ingunn Ms Engebretsen. 2011. "It Is Her Responsibility: Partner

Involvement in Prevention of Mother to Child Transmission of HIV Programmes, Northern Tanzania." *Journal of the International AIDS Society* 14:21.

Farquhar, Carey, James N. Kiarie, Barbra A. Richardson, Marjory N. Kabura, Francis N. John, Ruth W. Nduati, Dorothy A. Mbori-Ngacha, and Grace C. John-Stewart. 2004. "Antenatal Couple Counseling Increases Uptake of Interventions to Prevent HIV-1 Transmission." *Journal of Acquired Immune Deficiency Syndromes (1999)* 37(5):1620-26.

France Lert. 2017. *Seine-Saint-Denis, a Department committed to the end of AIDS in 2030*.

Greig, Alan, Michael Kimmel, and James Lang. 2000. "Men, Masculinities & Development: Broadening Our Work Towards Gender Equality".

Haile, Fisaha and Yemane Brhan. 2014. "Male Partner Involvements in PMTCT: A Cross Sectional Study, Mekelle, Northern Ethiopia." *BMC Pregnancy and Childbirth* 14:65.

HAS. 2009. *Dépistage de l'infection par le VIH en France : Stratégies et dispositif de dépistage. synthèse et recommandaitons*. Haute Autorité de Santé.

HAS. 2018. *Reassessment of the screening strategy for Chlamydia trachomatis infections. public health recommendation*. Haute Autorité de Santé.

Homsy, Jaco, Rachel King, Samuel S. Malamba, Christine Opio, Julius N. Kalamya, Jonathan Mermin, Alice Okallanyi, and John H. Obonyo. 2007. "The Need for Partner Consent Is a Main Reason for Opting out of Routine HIV Testing for Prevention of Mother-to-Child Transmission in a Rural Ugandan Hospital". *Journal of Acquired Immune Deficiency Syndromes (1999)* 44(3):366-69.

INSEE. 2019. *Population by sex, age and immigration status in 2016 - Seine-Saint Denis department*.

Kiarie, James N., Joan K. Kreiss, Barbra A. Richardson, and Grace C. John-Stewart. 2003. "Compliance with Antiretroviral Regimens to Prevent Perinatal HIV-1 Transmission in Kenya." *AIDS (London, England)* 17(1):65-71.

Kigozi, Isaac M., Loren M. Dobkin, Jeffrey N. Martin, Elvin H. Geng, Winnie Muyindike, Nneka I. Emenyonu, David R. Bangsberg, and Judith A. Hahn. 2009. "Late-Disease Stage at Presentation to an HIV Clinic in the Era of Free Antiretroviral Therapy in Sub-Saharan Africa." *Journal of Acquired Immune Deficiency Syndromes (1999)* 52(2):280-89.

Larsson, Elin C., Anna Thorson, Xavier Nsabagasani, Sarah Namusoko, Rebecca Popenoe, and Anna Mia Ekström. 2010. "Mistrust in Marriage--Reasons Why Men Do Not Accept Couple HIV Testing during Antenatal Care- a Qualitative Study in Eastern Uganda." *BMC Public Health* 10:769.

Marty, Lise, Françoise Cazein, Henri Panjo, Josiane Pillonel, Dominique Costagliola, Virginie Supervie, and HERMETIC Study Group. 2018. "Revealing Geographical and Population Heterogeneity in HIV Incidence, Undiagnosed HIV Prevalence and Time to Diagnosis to Improve Prevention and Care: Estimates for France." *Journal of the International AIDS Society* 21(3):e25100.

Mohlala, Boshishi K. F., Marie-Claude Boily, and Simon Gregson. 2011. "The Forgotten Half of the Equation: Randomized Controlled Trial of a Male Invitation to Attend Couple Voluntary Counselling and Testing". *AIDS (London, England)* 25(12):1535-41.

Nkuoh, Godlove N., Dorothy J. Meyer, Pius M. Tih, and Joseph Nkfusai. 2010. "Barriers to Men's Participation in Antenatal and Prevention of Mother-to-Child HIV Transmission Care in Cameroon, Africa". *Journal of Midwifery & Women's Health* 55(4):363-69.

Op de Coul, Eline L. M., Ard van Sighem, Kees Brinkman, Birgit H. van Benthem, Marchina E. van der Ende, Suzanne Geerlings, Peter Reiss, and ATHENA National Observational HIV Cohort. 2016. "Factors Associated with Presenting Late or with Advanced HIV Disease in the Netherlands, 1996-2014: Results from a National Observational Cohort". *BMJ Open* 6(1):e009688.

Orne-Gliemann, Joanna, Eric Balestre, Patrice Tchendjou, Marija Miric, Shrinivas Darak, Maia Butsashvili, Eddy Perez-Then, Fred Eboko, Melanie Plazy, Sanjeevani Kulkarni, Annabel Desgrées du Loû, François Dabis, and Prenahtest ANRS 12127 Study Group. 2013. "Increasing HIV Testing among Male Partners". *AIDS (London, England)* 27(7):1167-77.

Osoti, Alfred Onyango, Grace John-Stewart, James Njogu Kiarie, Richardson Barbra, John Kinuthia, Daisy Krakowiak, and Carey Farquhar. 2015. "Home-Based HIV Testing for Men Preferred over Clinic-Based Testing by Pregnant Women and Their Male Partners, a Nested Cross-Sectional Study." *BMC Infectious Diseases* 15:298.

PEPFAR. 2010. *Prevention of Mother-to-Child Transmission of HIV: Expert Panel Report and Recommendations to the U.S. Congress and U.S. Global AIDS Coordinator*.

Piffaretti, Clara, Vincent Looten, Sylvie Rey, Jeanne Fresson, Anne Fagot-Campagna, and Philippe Tuppin. 2018. "Management of Pregnancy Based on Healthcare Consumption of Women Who Delivered in France in 2015: Contribution of the National Health Data System (SNDS)". *Journal of Gynecology Obstetrics and Human Reproduction* 47(7):299-307.

Santé Publique France. 2019. "HIV testing and discoveries in Paris: 2018 epidemiological update".

Semrau, Katherine, Louise Kuhn, Cheswa Vwalika, Prisca Kasonde, Moses Sinkala, Chipepo Kankasa, Erin Shutes, Grace Aldrovandi, and Donald M. Thea. 2005. "Women in Couples Antenatal HIV Counseling and Testing Are Not More Likely to Report Adverse Social Events". *AIDS (London, England)* 19(6):603-9.

Thomson, Kerry A., James Hughes, Jared M. Baeten, Grace John-Stewart, Connie Celum, Craig R. Cohen, Kenneth Ngure, James Kiarie, Nelly Mugo, Renee Heffron, and Partners in Prevention HSV/HIV Transmission Study and Partners PrEP Study Teams. 2018. "Increased Risk of HIV Acquisition Among Women Throughout Pregnancy and During the Postpartum Period: A Prospective Per-Coital-Act Analysis Among Women With HIV-Infected Partners." *The Journal of Infectious Diseases* 218(1):16-25.

Wertz, Jocelyn, Jason Cesario, Jennifer Sackrison, Sean Kim, and Chi Dola. 2011. "Acute HIV Infection in Pregnancy: The Case for Third Trimester Rescreening". *Case Reports in Infectious Diseases* 2011:340817.

Word Health Organization. 2010. "PMTCT Strategic Vision 2010-2015: Preventing Mother to Child Transmission of HIV to Reach the UNGASS and MIllenium Development Goals Geneva 2010." Accessed August 2, 2019 (https://www.who.int/hiv/pub/mtct/strategic_vision/en/).

**List of annexes attached to the protocol :**

**Appendix 1 - Information note to participants, opt-out form**

**Appendix 2 - Maternal questionnaire**

**Appendix 3 - Paternal questionnaire**

**Appendix 4 - Refusal questionnaire**

**Appendix 5 - Favourable opinion of the CPP**

**Appendix 6 - CNIL agreement**

**Appendix 7 - Communication materials**
