## Supplementary File 2 for "Does a prenatal consultation dedicated to fathers’ health widen men’s access to prevention and care? A monocentric interventional research in the Paris metropolitan area"

**3038** **women gave a number allowing to call a father living in Paris metropolitan area, or the father was present at mother inclusion**

3038 eligible men were called or met the interviewers in the maternity ward, at mother inclusion

**522** never picked up or called back

**2516 eligible fathers were actually contacted**

**5** oppositions to research

**33** refused consultation at MQ

**345** refused by phone or e-mail

**366** never scheduled an appointment

**434** never attended sheduled appointments

**1333** eligible fathers attended consultation

**14 extra fathers for which their partner had not given their contact details or responded to the maternity questionnaire**, **or who lived out of Paris metropolitan area, made an appointment**

.

**1347** fathers attended consultation

Supplementary File 2: Simplified study flow-chart, Montreuil, 2021-2022
