## Supplementary File 3 for "Does a prenatal consultation dedicated to fathers’ health widen men’s access to prevention and care? A monocentric interventional research in the Paris metropolitan area"

**Appendix 1: Father Questionnaire** - imbedded in the Paternal Prenatal Consultation (PPC) frame

**The medical doctor or midwife first presents the project and collects consent:**

In Montreuil, we try to take an interest in the good health of men too, because the health of the family depends on the health of both parents. That's why we offer a prenatal consultation to all men who are going to have a child in our hospital. I am (identity, occupation), and together we will discuss your health, vaccinations and infectious disease status, and check that you have easy access to the health professionals you need. Afterwards, I will take your blood pressure, check your vaccinations and suggest a biological check-up.

All information concerning you is confidential, and will not be passed on to the future mother or to the maternity team.

We are conducting a study among fathers to better understand their health and prevention needs, before extending this consultation to all men who are about to have a child. Do you agree that I may ask you some questions, not all of which correspond to your situation, and that your answers may be used, anonymously, to improve our knowledge of the health of future fathers? This will take us about 10 minutes.

**Do you have any other children, already born?**

Yes, with the same mother -> how many

Yes, with another mother -> how many

No

*If the participant is born abroad:*

Number of children living abroad

Number of children living in France

*If children already born:*

Did you have any screening or consultation during your last pregnancy?

-> If yes, was it as part of Montreuil fathers' screening project (pilot research)? Alternatively, in which context?

**Who first told you about the paternal prenatal consultation?**

The expectant mother, the reception agent opening the prenatal file, the reception agent at a prenatal consultation, a health professional at the hospital, a health professional outside the hospital, a member of the research team at the Maternity Ward, a member of the research team by e-mail, a member of the research team by telephone, a friend/relative, through media (poster/leaflet/emission/article), an association, other (+specification)

**How did you make the appointment?**

I came with no appointment, my partner made the appointment for me, the research team suggested the appointment, I made the request, I don't know

Method for booking: e-mail, Doctolib®, telephone, face-to-face, I don't know, other (+specification)

**In which country were you born?**

*If born abroad:*

In what year did you arrive in France?

What was your main reason for coming to France?

Seek a better life, find work / Join my partner / Join another member of my family / Threats in my homeland / Studies / Medial reasons / Other (+specification)

**What is your current administrative situation?** *Or: are you legally resident? Irregular?*

*Then the investigator ask for details:*

No papers at all, Short-stay permit (Provisional residence permit/< 1 year Residence permit), 1-9 years residence permit, 10 years resident card, French ID, European Union (EU) ID or EU residence permit

**Do you have health insurance?** *Or what do you currently have to cover your health expenses?*

Complete Health Insurance, Basic Health Insurance only, State Medical Assistance (“Aide Médicale de l’Etat in French), foreign insurance, European Union health insurance card, nothing

**What is your job**: free text

**What is your current employment situation?**

Unemployed or non-contracted job, Precarious fixed-term contract or part-time work or temporary work or precarious temporary work or precarious self-employed, Permanent contract or non-precarious self-employment or non-precarious fixed-term contract or non-precarious temporary work, Student or in training

**How are you currently housed?**

In personal accommodation (you live at home, renting or owning your own home), Hosted by family or acquaintances, In a hostel or in a center for asylum seekers, Sheltered by associations, Sleeping rough or calling every day the social emergency number for overnight shelter

**Did you go to school? If you went to school, what was the last class you attended?**

Never, Primary or Koranic school, Secondary, Higher education

**How long have you been a couple?**

<1 year, 1-3 years, 4-10 years, > 10 years, we're not really together

**Do you live together?**

Yes, no, more or less (possible specification if more or less

**Are you married?**

No, Yes monogamous (any formal union: traditional, civil, religious), Yes polygamous or other concurrent household

**What is the household's main source of income?**

Me, my partner, roughly balanced, no income other than welfare benefits, if any

**In the last 12 months, have you had more than one sexual partner?** (Including the future mother)

Yes,No ; If yes : Female(s) : yes/no, Male(s) : yes/no, Occasional yes/no, Regular yes/no

**How often and in what context do you have contact with the healthcare system?**

1- I have a GP and I can see him/her if I need to, but it's not regular
2- I don't have a GP and very rarely come into contact with the care system
3- I have a doctor (GP or specialist) whom I see regularly for a chronic condition
4- I haven't seen anyone since I became an adult (except possibly at work),
5- I haven't seen anyone since I arrived in France (for immigrants)
6- Since I have been an adult (or since I arrived in France), I have only seen physicians in hospital emergency rooms

If answer 1 to 3**: When was your last medical appointment in France?** (approximate year)

**Have you ever had a HIV test**? If yes: **date of last test** (year)/**result** (negative, positive, doubtful)

*If seropositive* (= positive result to previous question): Are you being monitored for HIV?” (yes and on antiretroviral treatment and last consultation < 1 year / previous monitoring but no consultation for > 1 year or no treatment or treatment breakthrough / never monitored. Is your viral load undetectable? (Yes, No, Don't know). Have you discussed this with the child's mother? Yes, no

*If “yes” to the test question*:

**What was the reason for the last screening?**

My partner's pregnancy, On my own initiative or at my partner's request, offered by a healthcare provider, offered by an association/collective screening/street screening...

**Have you ever done a test with the mother of your unborn child, or exchanged the results of tests done separately?**

Yes (joint test or exchange of results), No, but we have talked about testing, No and we never talked about it, I did a test and gave her the results, She did a test and gave me the results

**Do you know about your hepatitis B status?**

I had it and am cured**,** I have chronic hepatitis B and am being monitored**;** I have chronic hepatitis B and I am not being monitored**,** I have been vaccinated**,** I was screened and told I was protected**,** I have been screened and only told I was negative**,** I don't know what it is or what my status is

*For the investigator, look at the vaccination chart and biological tests if any to complete*

**Have you ever been tested for hepatitis C?**

Yes, no, I don't know

*If born in France*: Have you received any vaccinations > 18? Yes, No, I don't know

**Have you ever had a sexually transmitted infection?** Yes, No, I don't know

*If yes*, **do you remember which one?** (Drop-down menu: hepatitis C is included in STIs, even if other modes of transmission are involved)

**Have you been the victim of violence? (**Other than an occasional brawl or altercation): Yes/no

*If yes*: physical violence, sexual violence, both + specific questions related to violence

*Clinical consultation. At the end of PPC:*

**What persuaded you to come to the PPC?**

**Do you think this consultation is adapted to the health needs of future fathers?** Suggestions for improving its content - free text, no limit.

**What can be done to ensure that as many fathers as possible accept it?** Suggestions for improving the format (location, schedule, content)
