## Supplementary File 4 for "Does a prenatal consultation dedicated to fathers’ health widen men’s access to prevention and care? A monocentric interventional research in the Paris metropolitan area"

**Supplementary file 4: Procedures for Paternal Prenatal Consultation (PPC) and biological tests**

The medical doctor or midwife explains PPC’s goal and process, collects consent, measures blood pressure, proceeds with the father’s questionnaire, provides a clinical examination if symptoms are present, checks vaccination, prescribes biological tests.

Blood sample is collected on site immediately after the consultation by a nurse, medical doctor or midwife

**HIV test:** All participants (except for documented HIV+ status and for participants tested during or just prior to the current pregnancy) – by Abbott ELISA serology or INSTI Rapid Antibody Assessment

**Anti HBs antibodies**: Participants born in France with no proven immunity (complete vaccination or previous >= 100 mUI/mL antibodies) except for known Ag HBS+ status

**AntiHBs antibodies+AntiHBc antibodies+HBs antigen**: participants born abroad with no proven immunity and no documented Ag HBs+; participants first tests with Anti HBs antibodies with a < 100mUI/mL (automatic addition)

**Hepatitis C serology**: participants born abroad with no history of Hepatitis C; participants born in France with history of blood exposure

**Hepatitis C viral load**: participants with history of hepatitis C and no negative viral load documented

**Urine molecular amplification test for *C. trachomatis* and *N. gonorrhoea****:* < 30 years participants and participants with 2 or more sexual partners during past 12 months

**Bilharziasis serology:** participants born or grown up in endemic area, when never tested before

**Glycemia:** participants at risk of undiagnosed diabetes, with symptoms of diabetes (polyuria polydipsia) or with lost to follow-up diabetes

**Any other tests depending on clinical presentation and medical history**

**Biological analysis results** are delivered and explained face-to-face, by telephone or by e-mail, depending on participant’s choice in the case of normal results, and face-to-face if anomalies are detected.

Vaccination update are prescribed immediately if vaccination chart is brought or at a second consultation if the vaccination chart (or pictures) is not brought but accessible. Vaccination up-date is offered immediately in the absence of complete health insurance coverage and if bringing back vaccines or updating them elsewhere would be difficult for the participant.

Participants are referred (with a letter and appointment booking if required) according to the needs
