## Supplementary File 5 for "Does a prenatal consultation dedicated to fathers’ health widen men’s access to prevention and care? A monocentric interventional research in the Paris metropolitan area"

**Supplementary file 6: Detailed results for HIV screening, diagnosis and referrals (N= 1347) during paternal prenatal consultation (PPC), Montreuil, 2021-2022**

1- HIV screening

|  | n/N | % |
| --- | --- | --- |
| Test following PPC | 1276/1347 | 94.7% |
| Tested during current pregnancy or as part of Medically Assisted Procreation procedure | 50/1347 | 3.7% |
| Refusal | 15/1347 | 1.1% |
| Prescription error or hemolyzed tube | 5/1347 | 0.4% |
| Known HIV-positive status | 1/1347 | 0.1% |

2- Diagnosis and pathologies remitted into care

|  | n |
| --- | --- |
| **Infectious disease (excluding dermatological infections)** | **106** |
| Incident Hepatitis B | 35 |
| Lost to follow up Hepatitis B | 18 |
| Hepatitis C (unknown positive serology) | 5 |
| Syphilis (unknown positive treponemal and nontreponemal tests) | 4 |
| *C.trachomatis* urethral carriage | 9 |
| *N. gonorrhoea* urethral carriage | 1 |
| Bilharziasis (unknown positive serology) | 31 |
| **Cardio-metabolic** | **63** |
| High Blood Pressure | 45 |
| Diabetes/glucose intolerance | 8 |
| Metabolic syndrome | 7 |
| **Dermatological** | **28** |
| **Psychological** | 17 |
| **Neuromuscular, rheumatological** | 16 |
| **Gastrointestinal and proctological** | 12 |
| **Respiratory, ear, nose and throat** | 12 |

3- Healthcare and social referrals

|  | n |
| --- | --- |
| New General Practitioner (GP) | 64 |
| Redirected to the attending physician for care | 42 |
| Free Hospital Unit for patients with no healthcare coverage | 26 |
| Hospital Specialist Physician^1^ | 60 |
| Specialist physician outside the hospital^1^ | 9 |
| Ophthalmologist | 3 |
| Proctologist | 3 |
| Addictologist | 7 |
| Dental care specialist | 12 |
| Psychologist, sexologist | 34 |
| Physiotherapist, osteopath, podiatrist | 8 |
| HIV-PrEP consultation / effective HIV-PrEP follow-up | 3/1 |
| Urologist for Vasectomy | 2 |
| Midwife for individual preparation to parenthood | 3 |
| Social Worker | 110 |
| Health Mediator | 65 |

^1^ Sexologist, addictologist, ophthalmologist, proctologic and physician running HIV-PrEP consultations are excluded
