## Supplementary File 6 for "Does a prenatal consultation dedicated to fathers’ health widen men’s access to prevention and care? A monocentric interventional research in the Paris metropolitan area"

|  |  |  | Univariate regression^*^ |
| --- | --- | --- | --- |
|  |  |  | (N = 1347) |
|  | **n** | **%** | OR [IC 95%] |
| Overall participants | 145 | 11 % |  |
| Migration status |  |  |  |
| French-born | 3 | 0.6 % | 1.00 |
| Immigrant | 142 | 17 % | **34 [11-107]** |
| Region of birth |  |  |  |
| France | 3 | 0.6 % | 1.00 |
| North Africa-Middle East | 26 | 10 % | **20 [6-66]** |
| Sub-saharan Africa | 104 | 26 % | **60 [19-192]** |
| Asia | 4 | 4 % | **7.27 [1.60-33.04]** |
| Rest of Europe-America | 8 | 7 % | **13.5 [3.52-51.86]** |
| School level |  |  |  |
| University | 29 | 5 % | 1.00 |
| Secondary school | 62 | 10 % | **1.96 [1.24-3.09]** |
| None, primary or koranic school | 54 | 29 % | **7.03 [4.31-11.48]** |
| Age |  |  |  |
| < 35 | 62 | 9 % | 1.00 |
| 35 and over | 83 | 13 % | **1.44 [1.02-2.05]** |
| Administrative status |  |  |  |
| Regular situation | 28 | 2 % | 1.00 |
| No or < 1 year residence permit | 117 | 54 % | **45 [29-72]** |
| Length of stay (immigrants) |  |  |  |
| 7 years and over | 51 | 10 % | 1.00 |
| 3-7 years | 75 | 30 % | **3.99 [2.69-5.93]** |
| <= 2 years | 16 | 25 % | **3.10 [1.64-5.86]** |
| Employment status |  |  |  |
| Non-precarious employment | 32 | 3 % | 1.00 |
| Precarious employment | 72 | 30 % | **12.04 [7.69-18.84]** |
| Unemployment, education or training | 41 | 23 % | **8.53 [5.19-14.02]** |
| Health insurance coverage |  |  |  |
| Complete Health Insurance | 24 | 2 % | 1.00 |
| Basic Health Insurance | 12 | 8 % | **3.38 [1.66-6.92]** |
| State Medical Assistance (SMA) | 30 | 33 % | **19.84 [10.9-36.0]** |
| No | 79 | 72 % | **106 [59-190]** |
| Integration into the healthcare system |  |  |  |
| Primary care physician or chronic disease follow-up | 54 | 6 % | 1.00 |
| Rarely or never in contact with a physician | 91 | 25 % | **5.73 [3.99-8.23]** |

**Supplementary table 1**: factors associated with referral to at least one social professional, Montreuil, 2021-2022

**^*^** Multivariate regression is not shown, as explaining factors overlap referral criteria
