## Supplementary File 7 for "Does a prenatal consultation dedicated to fathers’ health widen men’s access to prevention and care? A monocentric interventional research in the Paris metropolitan area"

**Supplementary table 2: Univariate and multivariate logistic regression analysis of factors associated with any diagnosis or pathology brought back into care in immigrant participants, Montreuil, 2021-2022 (N=842)**

|  | Participants  (n/N) | Percentage of participants | Univariate analysis  OR [IC 95] N = 842 | Final multivariate model^a^  aOR [IC 95] N=842 |
| --- | --- | --- | --- | --- |
| Any diagnosis or pathology brought back into care among immigrant participants | 183/842 | 22% |  |  |
| **Region of birth** |  |  |  |  |
| North Africa-Middle East | 28/247 | 11 % | 1.00 | 1.00 |
| Sub-saharan Africa | 124/392 | 32 % | **3.62 [2.31-5.66]** | **3.22 [2.02-5.13]** |
| Asia | 16/96 | 17% | 1.56 [0.80-3.04] | 1.67 [0.85-3.29] |
| Rest of Europe-America | 15/107 | 14 % | 1.27 [0.65-2.50] | 1.13 [0.57-2.25] |
| **School level** |  |  |  |  |
| University | 45/273 | 16% | 1.00 | 1.00 |
| Secondary school | 84/391 | 21% | 1.39 [0.93-2.07] | 1.37 [0.90-2.09] |
| None, primary or koranic school | 54/178 | 30% | **2.21 [1.40-3.47]** | 1.47 [0.91-2.38] |
| **Age** |  |  |  |  |
| < 35 | 86/365 | 24% | 1.00 |  |
| 35 and over | 97/477 | 20% | 0.83 [0.60-1.15] |  |
| **Administrative status** |  |  |  |  |
| Regular situation | 107/624 | 17% | 1.00 |  |
| No or < 1 year residence permit | 76/218 | 35% | **2.59 [1.83-3.66]** |  |
| **Length of stay** |  |  |  |  |
| 7 years and over | 98/526 | 18 % | 1.00 |  |
| 3-7 years | 70/250 | 28 % | **1.70 [1.19-2.42]** |  |
| <= 2 years | 15/64 | 23 % | 1.34 [0.72-2.48] |  |
| **Employment status** |  |  |  |  |
| Non-precarious employment | 91/502 | 18% | 1.00 |  |
| Precarious employment | 57/199 | 29% | **1.81 [1.24-2.66]** |  |
| Unemployment or training | 35/141 | 25% | 1.49 [0.96-2.33] |  |
| **Health insurance coverage** |  |  |  |  |
| Complete Health Insurance | 97/526 | 18% | 1.00 | 1.00 |
| Basic Health Insurance | 19/116 | 16% | 0.87 [0.50-1.48] | 0.83 [0.48-1.45] |
| State Medical Assistance (SMA) | 22/91 | 24% | 1.41 [0.83-2.39] | 1.10 [0.63-1.91] |
| No | 45/109 | 41% | **3.11 [2.00-4.83]** | **2.61 [1.64-4.14]** |
| **Integration into the healthcare system** |  |  |  |  |
| Primary care physician or chronic disease follow-up | 106/576 | 18% | 1.00 |  |
| Rarely or never in contact with a physician | 77/266 | 29% | **1.81 [1.29-2.53]** |  |
| **^a^** Variables included in the complete multivariate model were: region of birth, length of stay, school level, age, employment status, health insurance coverage. Administrative status and integration into the healthcare system were excluded because of collinearity with health insurance coverage. | | | | |

|  | Participants  (n/N) | Percentage of participants | Univariate analysis  OR [IC 95] N = 842 | Final multivariate model^a^  aOR [IC 95] N=842 |
| --- | --- | --- | --- | --- |
| Immigrant participants referred to one or several healthcare professionals | 166/842 | 20 % |  |  |
| **Region of birth** |  |  |  |  |
| North Africa-Middle East | 25/247 | 10 % | 1.00 | 1.00 |
| Sub-saharan Africa | 108/392 | 28 % | **3.38 [2.11-5.40]** | **2.99 [1.83-4.90]** |
| Asia | 18/96 | 19% | **2.05 [1.06-3.96]** | **2.32 [1.18-4.58]** |
| Rest of Europe-America | 15/107 | 14% | 1.45 [0.73-2.87] | 1.25 [0.62-2.53] |
| **School level** |  |  |  |  |
| University | 38/273 | 14% | 1.00 | 1.00 |
| Secondary school | 81/391 | 21% | **1.62 [1.06-2.46]** | 1.48 [0.95-2.31] |
| None, primary or koranic school | 47/178 | 26% | **2.22 [1.38-3.58]** | 1.38 [0.83-2.30] |
| **Age** |  |  |  |  |
| < 35 | 82/365 | 22% | 1.00 |  |
| 35 and over | 84/477 | 18% | 0.74 [0.52-1.04] |  |
| **Administrative status** |  |  |  |  |
| Regular situation | 94/624 | 15% | 1.00 |  |
| No or < 1 year residence permit | 72/218 | 33% | **2.78 [1.94-3.97]** |  |
| **Length of stay** |  |  |  |  |
| 7 years and over | 84/526 | 16 % | 1.00 |  |
| 3-7 years | 65/250 | 26 % | **1.85 [1.28-2.67]** |  |
| <= 2 years | 17/64 | 27 % | **1.90 [1.04-3.47]** |  |
| **Employment status** |  |  |  |  |
| Non-precarious employment | 78/502 | 15% | 1.00 |  |
| Precarious employment | 54/199 | 27% | **2.02 [1.36-3.00]** |  |
| Unemployment or training | 34/141 | 24% | **1.73 [1.10-2.72]** |  |
| **Health insurance coverage** |  |  |  |  |
| Complete Health Insurance | 75/526 | 14% | 1.00 | 1.00 |
| Basic Health Insurance | 23/116 | 20% | 1.49 [0.89-2.50] | 1.43 [0.84-2.43] |
| State Medical Assistance (SMA) | 23/91 | 25% | **2.03 [1.19-3.46]** | 1.73 [0.99-3.03] |
| No | 45/109 | 41% | **4.23 [2.69-6.65]** | **3.75 [2.33-6.01]** |
| **Integration into the healthcare system** |  |  |  |  |
| Primary care physician or chronic disease follow-up | 72/576 | 12% | 1.00 |  |
| Rarely or never in contact with a physician | 94/266 | 35% | **3.83 [2.69-5.44]** |  |
| **^a^** Variables included in the complete multivariate model were: region of birth, length of stay, school level, age, employment status, health insurance coverage. Administrative status and integration into the healthcare system were excluded because of collinearity with health insurance coverage. | | | | |

**Supplementary table 3: Univariate and multivariate logistic regression analysis of factors associated with immigrant participants being referred to any healthcare professional, Montreuil, 2021-2022 (N=842)**

|  | Participants  (n/N) | Percentage of participants | Univariate analysis  OR [IC 95] N = 842 | Final multivariate model^a^  aOR [IC 95] N=842 |
| --- | --- | --- | --- | --- |
| Immigrant participants that received one or several vaccine updates | 407/842 | 48% |  |  |
| **Region of birth** |  |  |  |  |
| North Africa-Middle East | 111/247 | 45 % | 1.00 | 1.00 |
| Sub-saharan Africa | 226/392 | 58 % | **1.67 [1.21-2.30]** | 1.39 [0.99-1.96] |
| Asia | 48/96 | 50 % | 1.22 [0.76-1.96] | 1.27 [0.78-2.07] |
| Rest of Europe-America | 50/107 | 47 % | 1.07 [0.68-1.69] | 0.98 [0.61-1.56] |
| **School level** |  |  |  |  |
| University | 123/273 | 45% | 1.00 | 1.00 |
| Secondary school | 201/391 | 51% | 1.29 [0.95-1.76] | 1.18 [0.86-1.63] |
| None, primary or koranic school | 111/178 | 62% | **2.02 [1.37-2.97]** | **1.56 [1.04-2.35]** |
| **Age** |  |  |  |  |
| < 35 | 190/365 | 52% | 1.00 |  |
| 35 and over | 245/477 | 51% | 0.97 [0.74-1.28] |  |
| **Administrative status** |  |  |  |  |
| Regular situation | 288/624 | 46% | 1.00 |  |
| No or < 1 year residence permit | 147/218 | 67% | **2.41 [1.75-3.34]** |  |
| **Length of stay** |  |  |  |  |
| 7 years and over | 252/526 | 48 % | 1.00 |  |
| 3-7 years | 145/250 | 58 % | **1.50 [1.11-2.03]** |  |
| <= 2 years | 37/64 | 58 % | 1.49 [0.88-2.52] |  |
| **Employment status** |  |  |  |  |
| Non-precarious employment | 236/502 | 47% | 1.00 |  |
| Precarious employment | 123/199 | 62% | **1.82 [1.30-2.55]** |  |
| Unemployment or training | 76/141 | 54% | 1.32 [0.91-1.92] |  |
| **Health insurance coverage** |  |  |  |  |
| Complete Health Insurance | 237/526 | 45% | 1.00 | 1.00 |
| Basic Health Insurance | 63/116 | 54% | 1.45 [0.97-2.17] | 1.42 [0.94-2.13] |
| State Medical Assistance (SMA) | 55/91 | 60% | **1.86 [1.18-2.93]** | **1.65 [1.03-2.64]** |
| No | 80/109 | 73% | **3.36 [2.13-5.32]** | **3.08 [1.93-4.92]** |
| **Integration into the healthcare system** |  |  |  |  |
| Primary care physician or chronic disease follow-up | 267/576 | 46% | 1.00 |  |
| Rarely or never in contact with a physician | 168/266 | 63% | **1.98 [1.47-2.67]** |  |
| **^a^** Variables included in the complete multivariate model were: region of birth, length of stay, school level, age, employment status, health insurance coverage. Administrative status and integration into the healthcare system were excluded because of collinearity with health insurance coverage. | | | | |

**Supplementary table 4: Univariate and multivariate logistic regression analysis of factors associated with any vaccination update in immigrant participants, Montreuil, 2021-2022 (N=842)**
